# Association of road traffic noise exposure with dementia or cognitive impairment – a systematic review of longitudinal cohort studies

**DOI:** 10.1101/2025.09.11.25335584

**Authors:** Emil Basil Scaria, Nisha Dhanda

**Affiliations:** Department of Applied Health Sciences, College of Medicine and Health, University of Birmingham, Dubai, United Arab Emirates; Department of Applied Health Sciences, College of Medicine and Health, University of Birmingham, Edgbaston, United Kingdom

**Keywords:** noise, transportation noise, dementia, cognitive dysfunction

## Abstract

**Background:** Road traffic noise is a major public health concern that is associated with various cardiometabolic and neurological disorders. Dementia poses a significant health and socioeconomic burden. Existing systematic reviews have not explored the link between road traffic noise exposure alone, and risk of dementia or cognitive impairment. This review aims to identify and summarise available evidence linking road traffic noise exposure with risk of dementia or cognitive impairment.

**Methods:** MEDLINE, EMBASE, CINAHL Plus and GreenFile were searched for studies on road traffic noise exposure and the risk of dementia or cognitive impairment among adults from inception to July 2025 without restrictions on setting or geographical location. Studies were identified using strict inclusion and exclusion criteria. Two independent reviewers conducted screening, data extraction, and quality assessment. A narrative synthesis was conducted.

**Results:** 3296 studies were retrieved from the searches, of which 3264 were excluded and 32 underwent full text screening. 8 studies were narratively synthesised. Quality assessment of the studies revealed that they were good quality and only one study was prone to a high risk of bias. The findings suggest that adults exposed to high levels of road traffic noise exposure, particularly >50 dB, compared to those exposed to lower levels of road traffic noise are at higher risk of developing dementia or cognitive impairment.

**Conclusion:** There is a positive association between road traffic noise and dementia or cognitive decline. However, the amount of good-quality evidence is low and larger longitudinal studies using robust methods are needed. Such research could have significant implications on infrastructure planning and development of regulations to prevent adverse health effects due to road traffic noise.

## 1. Introduction

Noise is an acoustic phenomenon of excessive or loud sound (1). It is also referred to as noise pollution, which is characterised by environmental noise that is disruptive and disturbing (2).

Chronic exposure to excessive noise has been shown to have detrimental effects on human health (3, 4). Anthropogenic noise, the primary cause of noise pollution, also has ecological implications. In recent years, research has shed light on the impact of noise pollution on human health, particularly in children and older adults. This includes a range of health outcomes such as cognitive and sleep disorders, mental health issues, cardiovascular diseases, and metabolic dysfunction (5, 6). Hearing impairment is the most common adverse effect of noise pollution (7, 8).

Globally, dementia affects an estimated 55 million individuals, with projections reaching 78 million by 2030 (9, 10). The World Health Organization’s Burden of Disease from Environmental Noise report (2011) quantified over one million healthy life years lost annually due to environmental noise exposure, predominantly from road traffic sources (11). Road traffic noise is defined as the A-weighted equivalent continuous sound level (Leq) generated by vehicular sources including cars, heavy goods vehicles, and motorcycles which is measured or modelled at residential façades (12).

Traffic noise exposure initiates a cascade of physiological responses that may contribute to neurodegenerative processes. Chronic exposure to elevated A-weighted Leq activates the hypothalamic–pituitary–adrenal (HPA) axis, resulting in sustained cortisol release and oxidative stress (13, 14). Animal studies have shown that both air and noise pollution cause neurodegenerative changes that lead to neuroinflammation and oxidative stress (15–17). Noise-induced sleep fragmentation further exacerbates microglial activation and blood–brain barrier permeability, promoting neuroinflammation and accumulation of β-amyloid and tau proteins (18, 19).

Dementia is a neurocognitive disorder that impairs memory, executive function, and other cognitive domains, making routine activities challenging (20). It develops gradually over many years, with symptoms beginning up to 10-12 years before onset (21, 22). The cognitive abilities of affected individuals decline over time, leading to neuropsychiatric symptoms such as confusion, anxiety, depression, and apathy (23, 24). They struggle with daily activities and are at a heightened risk for debilitating conditions like urinary incontinence and hip fractures (25, 26), significantly impacting their quality of life and dependence on nursing care (27, 28). In contrast, cognitive decline refers to reductions in performance in specific cognitive domains (e.g., memory, attention), whereas dementia denotes a clinical syndrome of multi-domain impairment that interferes with autonomy (29). In 1990, there were approximately 20.2 million people living with dementia worldwide, which rose to 43.8 million individuals by 2016 (30).

Dementia is a complex disorder with various factors contributing to its development, including diet, lifestyle, physical activity, sleep, genetics, and cardiovascular disease (31, 32) While there are cognitive tests, psychiatric evaluations, and brain imaging to diagnose dementia, there is currently no cure for dementia, and symptoms are managed using medications and non-pharmacological interventions. Cholinesterase inhibitors and memantine are used to manage cognitive decline, while lifestyle changes and occupational therapies improve quality of life (33). According to a 2019 study (34), the median survival for patients with dementia is 5.2 years, leading to significant public health resources being spent on managing the condition, providing nursing care, and treating comorbidities. The global annual cost of dementia, which included direct medical costs, direct social sector costs, and costs of informal care, was estimated at USD 1313.4 billion in 2019 (35).

In England, transportation noise has been recognised as a significant burden of disease, with an estimated 97,000 DALYs lost due to road traffic noise (36). However, accurately measuring road traffic noise is challenging, as it is impractical to measure the noise emitted by each individual vehicle and measuring personal exposure is intrusive. The World Health Organization recommends using standard indices that measure road traffic noise using Leq, provides an average value for the total sound produced in an area during a specific period (37, 38). The Leq value over an entire day can be expressed as Lden, which includes penalties for evening and night-time noise. Leq during night-time may also be expressed as Lnight. The WHO guidelines recommend that road traffic noise (Lden) should remain below 53 dB and road traffic noise at night (Lnight) should remain below 45 dB to prevent adverse health outcomes. These values are below the normal conversational level of 60 dB (39). Thus, road traffic noise is not only an environmental nuisance but also a recognised public health hazard whereby the thresholds are frequently exceeded in urban areas worldwide.

Understanding the potential link between traffic noise and dementia is therefore critical for informing urban planning, transport infrastructure design, and noise mitigation policies, which could yield co-benefits for cardiovascular, mental, and cognitive health.

Previous research has examined the impact of road traffic noise on cognitive performance and dementia using both cross-sectional (40) and cohort (41, 42) studies. However, these studies have yielded inconclusive results and been complicated by confounding factors and mediators. Although there is evidence linking road traffic noise exposure to hearing loss and hearing loss to dementia, there is insufficient conclusive evidence linking road traffic noise exposure to dementia directly. A longitudinal ecological time-series study (43) suggests that short-term exposure to traffic noise may exacerbate symptoms and increase hospital admission rates for dementia. Additional research has explored the relationship between exposure to both air pollution and noise with dementia. A large cohort study conducted in Canada found that living near high-traffic roads increased the incidence of dementia, which could be attributed to exposure to air or noise pollution (44). A cross-sectional analysis of participants aged 50-80 in the Heinz Nixdorf Recall study found that long-term exposure to air pollution and traffic noise was associated with mild cognitive impairment (40), with the association between traffic noise and global cognitive scores only seen with high exposure to air pollution (45). A study in South Korea observed 11% higher odds of cognitive decline in people who reported exposure to noise pollution in their residential area (46).

To the best of our knowledge, there are only three systematic reviews that have examined the relationship between road traffic noise exposure and the risk of cognitive impairment or dementia in adults (47–49). One review (47) had multiple outcomes, which may have affected the quality of studies related to dementia and cognition. Additionally, it was commissioned by the UK Department for the Environment, Food and Rural Affairs, and therefore had a political agenda limited to the UK context. Another review (48) noted that the studies included had low methodological quality, which prevented adequate pooling of results. A third study (49) conducted a meta-analysis to investigate the dose-dependent relationship between noise exposure and dementia risk, but the inclusion of case-control and cross-sectional studies made it difficult to determine the temporal relationship between the exposure and outcome variables. Moreover, the studies included were noted to have high levels of recall and selection bias. Therefore, a systematic review that includes longitudinal cohort studies investigating the association between chronic exposure to road traffic noise and dementia/cognitive impairment risk as a sole outcome is needed to clarify the existing evidence base on this topic.

### 1.1 Confounders and mediators

Figure 1 is a directed acyclic graph (DAG) that showcases the potential causal pathways linking road traffic noise to dementia, as depicted in the existing literature, while also identifying potential confounders and mediators in this relationship (50). .

**Figure 1:**
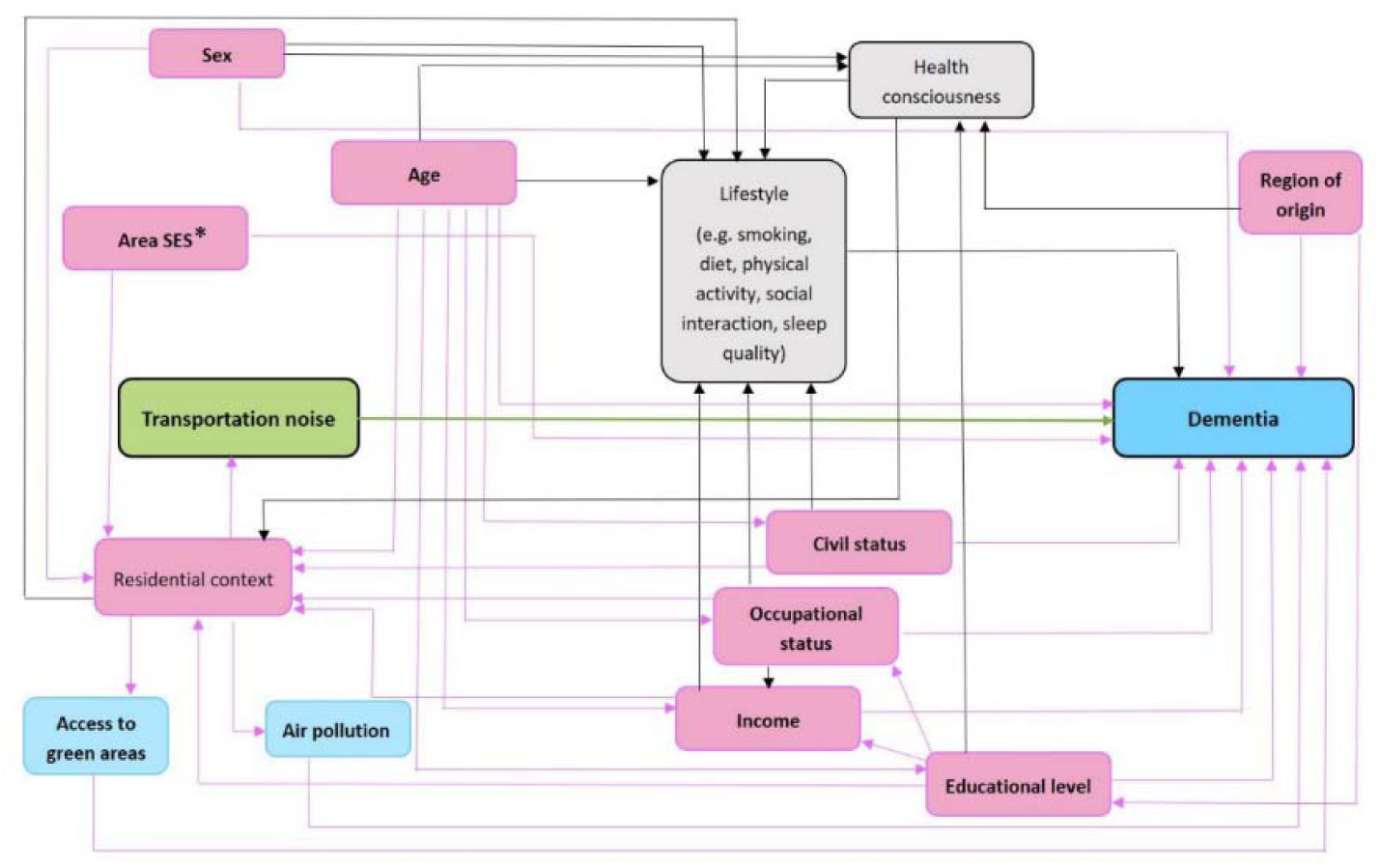
Directed Acyclic Graph showing the various mediators and potential pathways linking noise exposure with cognitive impairment and dementia. Reprinted from “Residential exposure to transportation noise in Denmark and incidence of dementia: national cohort study,” by M.L. Cantuaria et al., 2021, *BMJ*, 374, n1954. Copyright 2021 by BMJ Publishing Group Ltd. Reprinted with permission (50). *SES = socioeconomic status*.

Advancing age is the primary risk factor for dementia (51, 52), with women being at a slightly higher risk than men (53). Environmental noise exposure has been found to be higher for people of lower socioeconomic status (54, 55), and low socioeconomic status has been associated with an increased risk for developing dementia (56). Residential areas with higher levels of noise, such as high-rise buildings in urban areas, can also contribute to an increased risk of dementia (57). Social isolation, often a result of hearing loss caused by noise exposure (58), is another significant factor which has been linked to a higher likelihood of dementia (59).

Exposure to road noise is a significant public health issue that has significant impacts on individuals’ health and places a heavy burden on society (60). This review aims to systematically search, identify, and summarise the available evidence linking road traffic noise exposure with dementia or cognitive impairment, and to synthesise the findings narratively. A quantitative meta-analysis was planned but was not conducted due to substantial heterogeneity in study methods and outcomes.

## 2. Methods

The review was conducted following the Preferred Reporting Items for Systematic Reviews and Meta-analyses (PRISMA) guidelines (61).

### 2.1 Search Strategy

A scoping search and assessment of previous systematic reviews were carried out and used to design a search strategy. Systematic searches were carried out using tailored strategies on four databases - MEDLINE, EMBASE, CINAHL Plus and GreenFile in July 2025.

MEDLINE and EMBASE were searched as they are general bibliographic databases with broad coverage of biomedical and scientific literature. CINAHL Plus, with its extensive nursing and allied-health journal indexing, was searched to capture relevant clinical and epidemiologic studies; and GreenFILE was included as a dedicated environmental database covering different types of pollution and its human impact. A search for grey literature was conducted on BASE (Bielefeld Academic Search Engine) without imposing restrictions and using relevant keywords such as ‘road traffic noise’ and ‘dementia’. There were 43 results but no new results apart from those already considered for inclusion from the main searches were retrieved.

The search strategy included relevant search terms and MeSH headings. It was broad and included free text terms and MeSH headings related to traffic noise, noise pollution, dementia, cognitive decline, and cognitive impairment. They were combined appropriately using Boolean operators (AND, OR, NOT). Searches were not limited by time or language. The databases were searched from inception to July 2025. The strategy used for each database is available in Supplementary Materials (Supplementary Material A). The reference lists of published reviews and included studies were also searched to identify eligible studies. Full text articles for all studies were obtained using University of Birmingham library resources or through open access journals.

### 2.2 Study selection

A population-exposure-comparator-outcomes-study design (PECOS) was used to create the inclusion and exclusion criteria (see Table 1). They are as follows:

**Table 1:** Inclusion and exclusion criteria for study eligibility.

| Inclusion Criteria | Exclusion Criteria |
| --- | --- |
| Prospective and retrospective cohort studies. | Randomised controlled trials, cluster randomised controlled trials, controlled before-and-after studies, non-randomised studies, case-control studies, cross-sectional studies and case series or reports |
| Human studies | Animal studies |
| Studies including adults from the general population | Studies including only children below the age of 18 years |
| Studies with exposures related to traffic noise | Noise related exposures not related to traffic noise |
| Studies with traffic noise measured or modelled objectively | Studies with traffic noise measured subjectively |
| Studies with dementia or cognitive impairment as outcome | Studies with outcomes unrelated to dementia or cognitive impairment |
| Studies using any standard, internationally accepted diagnostic criteria like DSM-5 (20) | Studies not mentioning diagnostic criteria for outcome measurement. |
| Any mention of dementia, cognitive impairment, Alzheimer's Disease, or neurocognitive disorders as outcomes due to traffic noise exposure. | Studies that only have cognitive tests as outcomes. |
| No language restrictions. | Studies including individuals with hearing problems. |
| No timing restrictions |  |
| No restrictions based on publication status |  |
| No restrictions based on setting or geographical location. |  |

#### 2.2.1 Study design

Prospective and retrospective cohort studies were considered for inclusion. Dementia is a disease of insidious onset and may be difficult to detect without long duration of follow-up. As there are no existing systematic reviews that focus solely on longitudinal study designs to explore this association and account for the temporal nature of this relationship, we have chosen to exclude other types.

Randomised controlled trials and cluster randomised controlled trials have been excluded as high levels of noise exposure can cause adverse health outcomes, and randomisation to long-term interventions exposing participants to noise poses ethical concerns. Cross sectional studies were excluded because they cannot establish the temporal sequence between traffic noise exposure and subsequent onset of dementia, leaving findings vulnerable to reverse causation and survivorship biases. In addition, single time point assessments of both exposure and outcome are prone to bias, as participants with early, subclinical cognitive impairment may selectively relocate to quieter environments or be less likely to participate in noise surveys, thereby distorting true associations between exposure and outcome. By restricting our review to longitudinal cohort studies, we ensure that noise exposure is measured prior to disease onset, bolster causal inference through clear temporality, and minimise bias from unmeasured confounders and selective participation.

#### 2.2.2 Population

Studies with adults from the general population were included. Studies that only included children (below 18 years) were excluded as the outcome of interest is not expected in children. Dementia is a longstanding disease that develops over a span of 20-30 years with symptoms appearing commonly in the middle-aged or senile population (62). Furthermore, scoping and literature reviews revealed that most primary studies focused on adults aged above 40 years. Studies including both children and adults were excluded as they may be prone to high levels of bias. Animal studies were excluded.

#### 2.2.3 Exposure

Studies in which participants were exposed to road traffic noise and the exposure was measured or modelled objectively were included. Objective measurements of noise refer to measurements made by instruments such as noise level meters and dosimeters or predicted using computer models that generate estimates of noise levels over a given geographical area. Studies of all follow-up durations were included. Self-reported noise, occupational noise, neighbourhood noise, construction noise, or any other noise exposures unrelated to road traffic noise were excluded. Studies that measured noise exposure subjectively were also excluded as they may be prone to a high risk of bias.

#### 2.2.4 Comparator

Studies in which participants had no exposure or low levels of exposure to road traffic noise were the comparators. The participants with high noise exposure and those with low noise exposure could then be followed to detect the outcome.

#### 2.2.5 Outcomes

The primary outcomes were dementia or cognitive impairment. Studies using standard, internationally accepted diagnostic criteria such as Diagnostic and Statistical Manual of Mental Disorders, Fifth Edition (DSM-5) (20) were included. Studies that had only had cognitive tests as outcomes and studies including individuals with hearing problems were excluded.

#### 2.2.6 Timing

There were no restrictions based on timing.

#### 2.2.7 Setting

There were no restrictions based on setting or geographical location.

#### 2.2.8 Language

There were no restrictions based on language of publication. Studies in languages other than English would be translated with the help of a native speaker of that language from within the university. A study would be excluded only if such support was not available as obtaining a high-quality translation through other means would be time-consuming and resource intensive.

#### 2.2.9 Publication

There were no restrictions based on the publication date. Retrieved search results included studies from inception (1946 for MEDLINE; 1974 for Embase) to July 2025.

### 2.3 Screening

All studies were imported into EndNote 20.2.1, a reference management software by Microsoft (63), and duplicates were removed. They were uploaded into Covidence, a systematic review management software, which was used to screen abstracts and full texts, as well as resolve disagreements (64).

Title and abstract screening were performed by two independent reviewers (ES and ND) and articles that met the inclusion criteria were selected. Disagreements were resolved thorough discussion by ES and ND. The full texts of studies that were selected after title and abstract screening were screened independently by two reviewers (ES and ND) for inclusion in the review using Covidence.

Disagreements were resolved through discussion and review by ES and ND.

### 2.4 Data Extraction

A data extraction form was used to extract relevant information from the included studies. The form created using Microsoft Excel has been provided (see Supplementary Material B). Data extraction was conducted by the primary reviewer (ES) and verified by the second reviewer (ND). The form was piloted by one reviewer (ES) on two studies before further extraction was undertaken. A data extraction table was created to report the information extracted and help with comparison of extracted data. General information for each study such as title, authors, year of publication, study design, and location were extracted along with data related to population, exposure, outcome, and comparators. This included data related to noise source, exposure levels, measurement method, type of dementia, and diagnostic criteria, statistical methods, unadjusted and adjusted risk estimates such as hazard ratios (HR), 95% confidence intervals, p-values, confounders and covariates. Items that were used to conduct quality assessment were also extracted at this stage.

### 2.5 Outcomes

The primary outcomes were dementia or cognitive impairment. There are standard, internationally accepted diagnostic criteria for both outcomes such as DSM-5 and ICD10 codes. DSM-5 (20) is the most frequently used. Other diagnostic criteria are generally adapted from DSM-5 and follow similar standards for diagnosis. Diagnostic and Statistical Manual of Mental Disorders, Fourth Edition (DSM 4) (65) and International Classification of Diseases – 10^th^ Revision (ICD 10) (66, 67) are the most commonly used. All tools used to measure these outcomes along with any relevant criteria are summarised in Table 4.

### 2.6 Quality Assessment

The risk of bias was evaluated using the Risk of Bias In Non-randomised Studies–of Exposure (ROBINS-E) tool (68). It is a structured tool developed at the University of Bristol to assess the risk of bias in observational studies, especially for systematic reviews. It uses seven domains with signalling questions to thoroughly assess the risk of bias, and uses this to grade the overall risk of bias. There are some limitations to the tool such as its inability to distinguish between confounders and co-exposures, as well as the usage of a rating system that fails to differentiate between studies with single and multiple risks of bias (69). However, there is no perfect tool to assess the quality of observational studies of exposure, and ROBINS-E was considered more comprehensive and specific for this review compared to conventional quality assessment tools. Quality assessment items were extracted at the data extraction stage and two independent reviewers (ES and ND) conducted this process. Disagreements were resolved by discussion and review by ES and ND. The tool was piloted on an included study to assess its suitability before further steps were undertaken..

### 2.7 Synthesis

A meta-analysis was planned to pool effect estimates from eligible cohort studies where sufficient homogeneity in exposure metrics, outcome definitions, and effect measures existed. However, the included studies demonstrated substantial heterogeneity in noise metrics (Lden, Lnight, Leq), exposure categorisation, outcome ascertainment (all-cause dementia, Alzheimer’s disease, cognitive decline), and covariate adjustment. These differences precluded a meaningful pooled quantitative estimate. Therefore, a narrative synthesis was undertaken, structured to describe study characteristics, compare effect patterns, and explore reasons for heterogeneity. All extracted summary estimates are given in the Supplementary Materials (see Table C).

#### 2.7.1 Narrative synthesis

The included studies were synthesised narratively, detailing the findings and results in the studies. The results were tabulated in Table 4 to allow for comparison between the studies. The conclusions were summarised and overall themes or patterns in the inferences noted.

## 3. Results

### 3.1 Study screening and selection

The screening and selection process is summarised using a PRISMA diagram (Figure 2).

**Figure 2:**
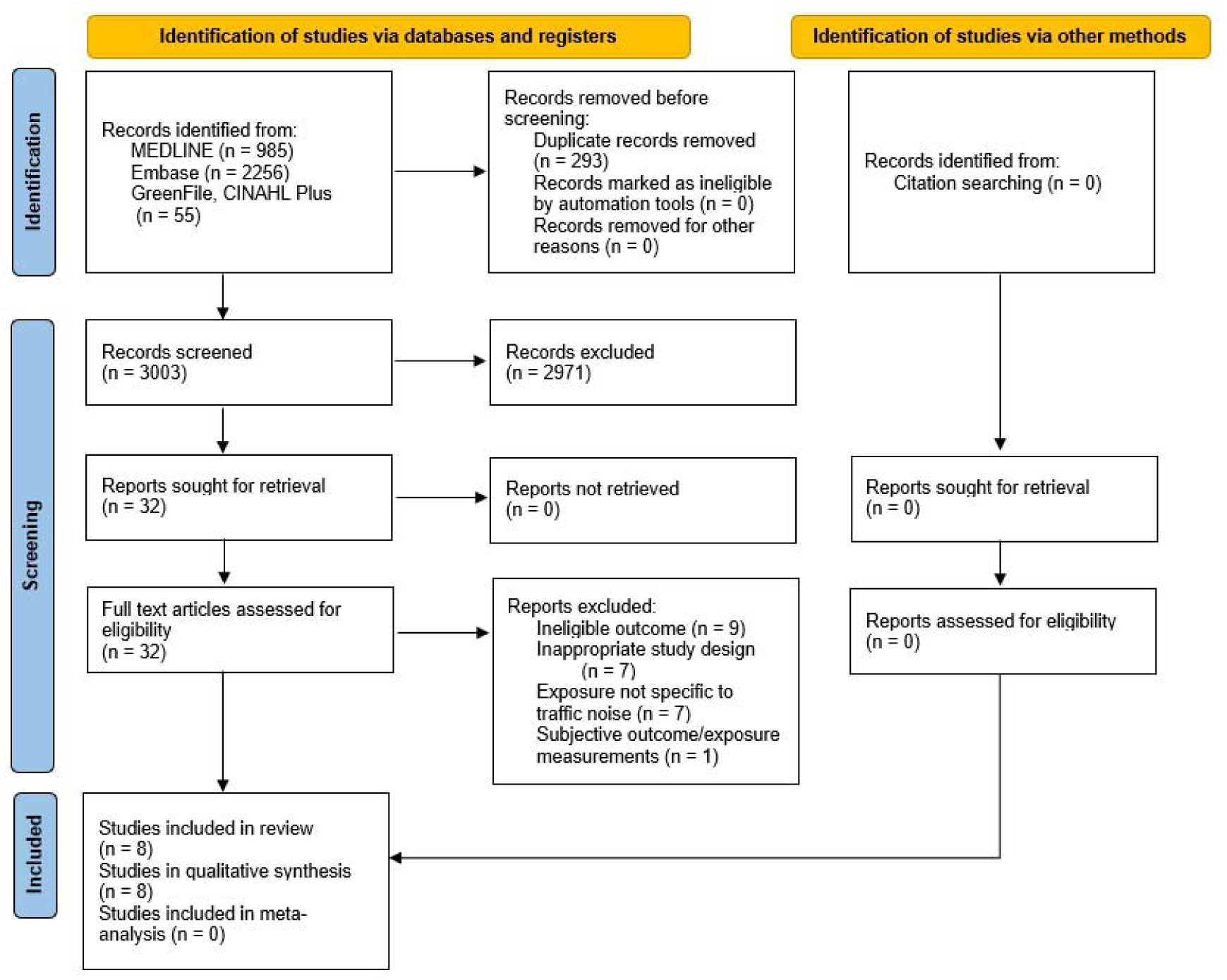
PRISMA (61) flowchart illustrating the study selection process.

A comprehensive search of the four databases (MEDLINE, Embase, CINAHL Plus, GreenFile) identified 3296 records. No new papers were identified by searching the reference lists of systematic reviews or included studies. After removal of duplicates (n = 293), there were 3003 studies that underwent title and abstract screening. 2971 studies excluded based on their irrelevance to the review question and their eligibility as per the inclusion criteria. Full texts of the remaining 32 studies were obtained. All the studies were in English and did not require translation. The inclusion and exclusion criteria were applied and resulted in 24 studies being excluded.

8 studies (50, 70–76) were selected for narrative synthesis, and the full texts and supplementary material were obtained. All the included studies used a cohort study design to explore the association between road traffic noise exposure and risk of dementia or cognitive impairment.

### 3.2 Study characteristics

Study characteristics are summarised in Table 2. All the studies included adults over the age of 40. Three studies (50, 73, 76) included adults aged over 60 years. One study only included female participants (75). The number of male and female participants in the other included studies was almost equal, except two studies. One study (73) had 58% female and 42% male participants, while the other (76) had 61% female and 39% male participants. Two studies (50, 70) did not have any missing participants and analysis included all the participants in the cohort. The remaining studies had to exclude participants from the analysis for various reasons, as detailed in Table 2. However, these studies conducted a complete case analysis and included only participants with complete data on exposure, outcome, and covariates in their analyses. The studies included a total of 2,571,591 participants, of which the overwhelming majority (n = 1,938,994) were from one study (50).

**Table 2:**
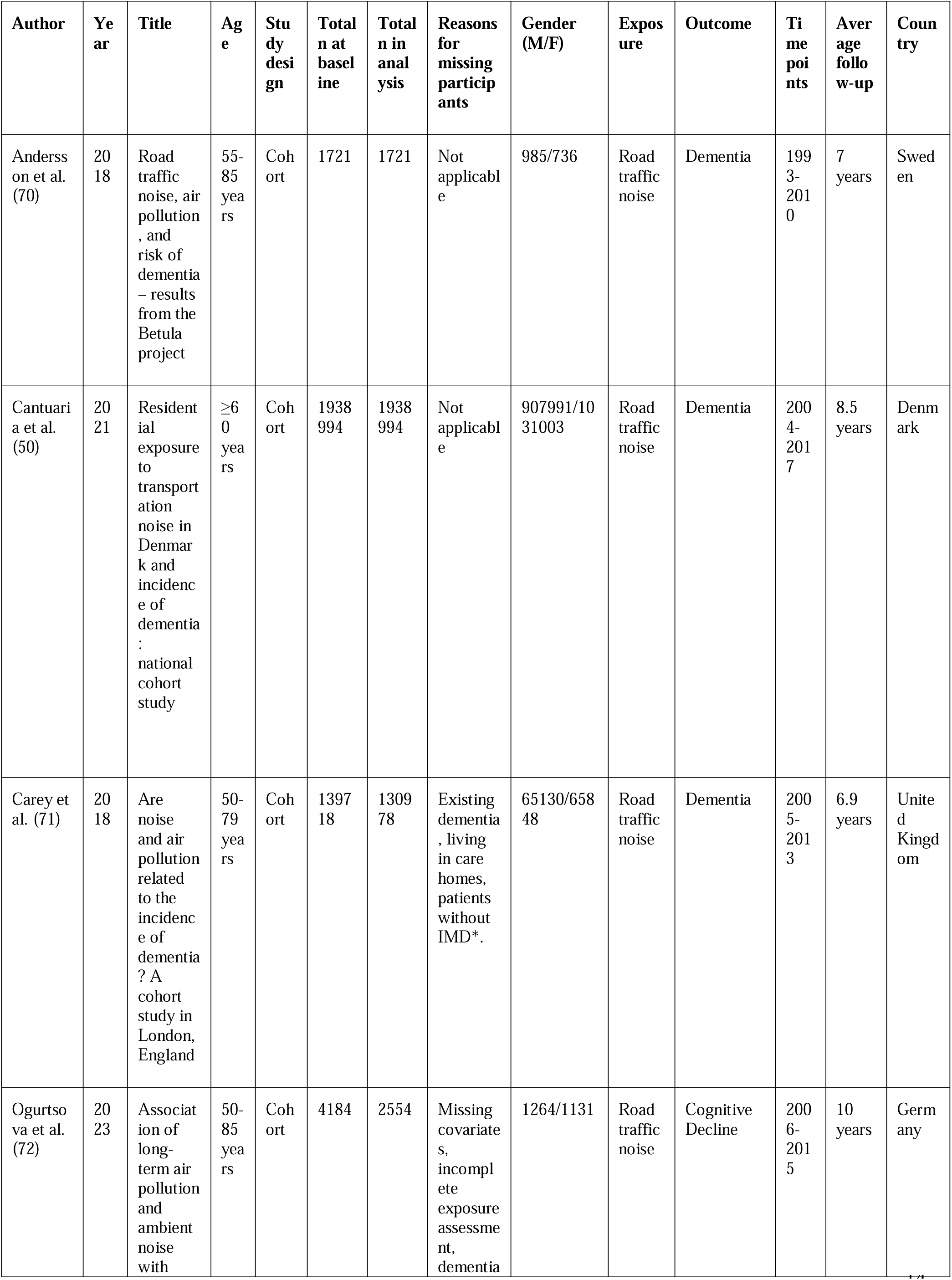
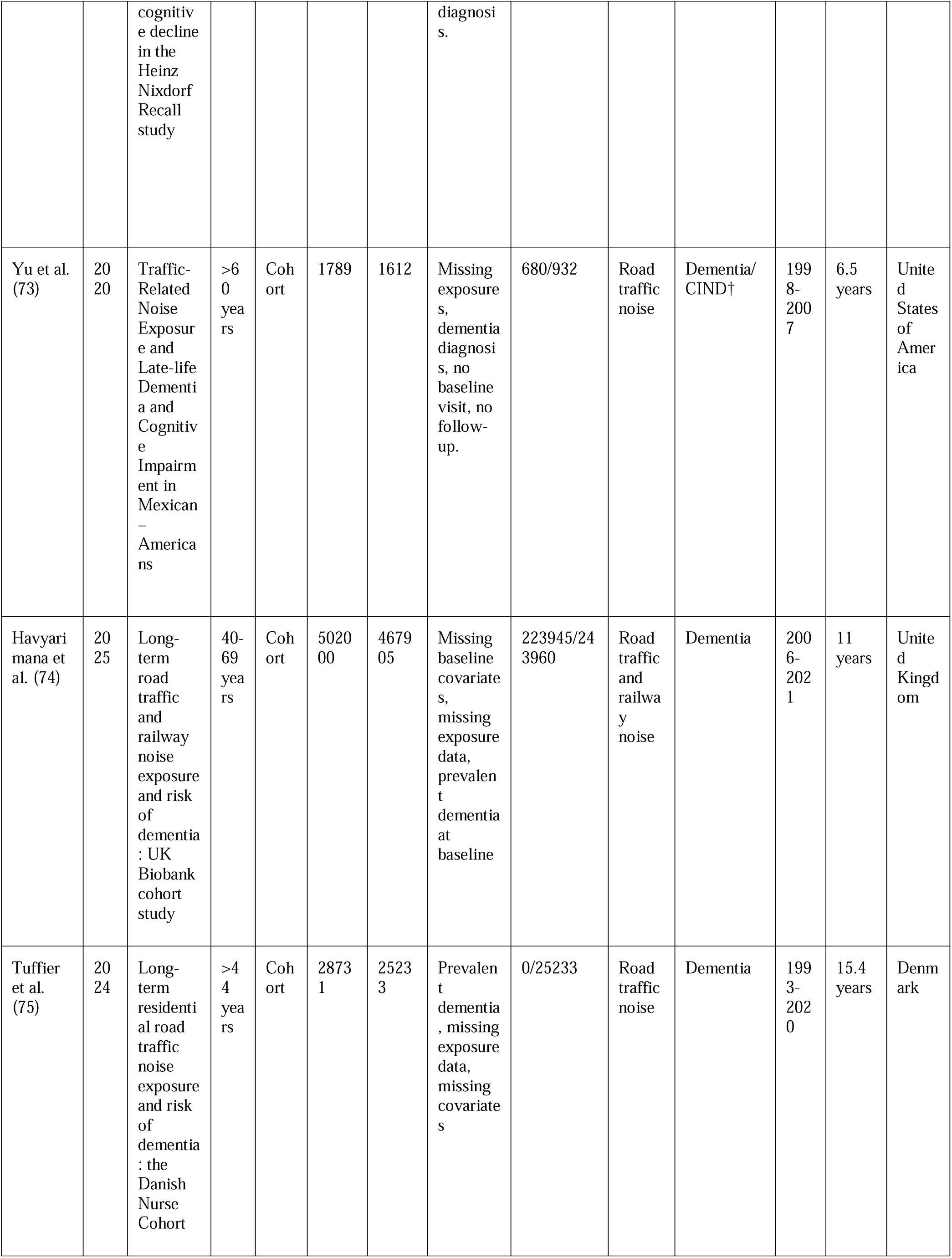
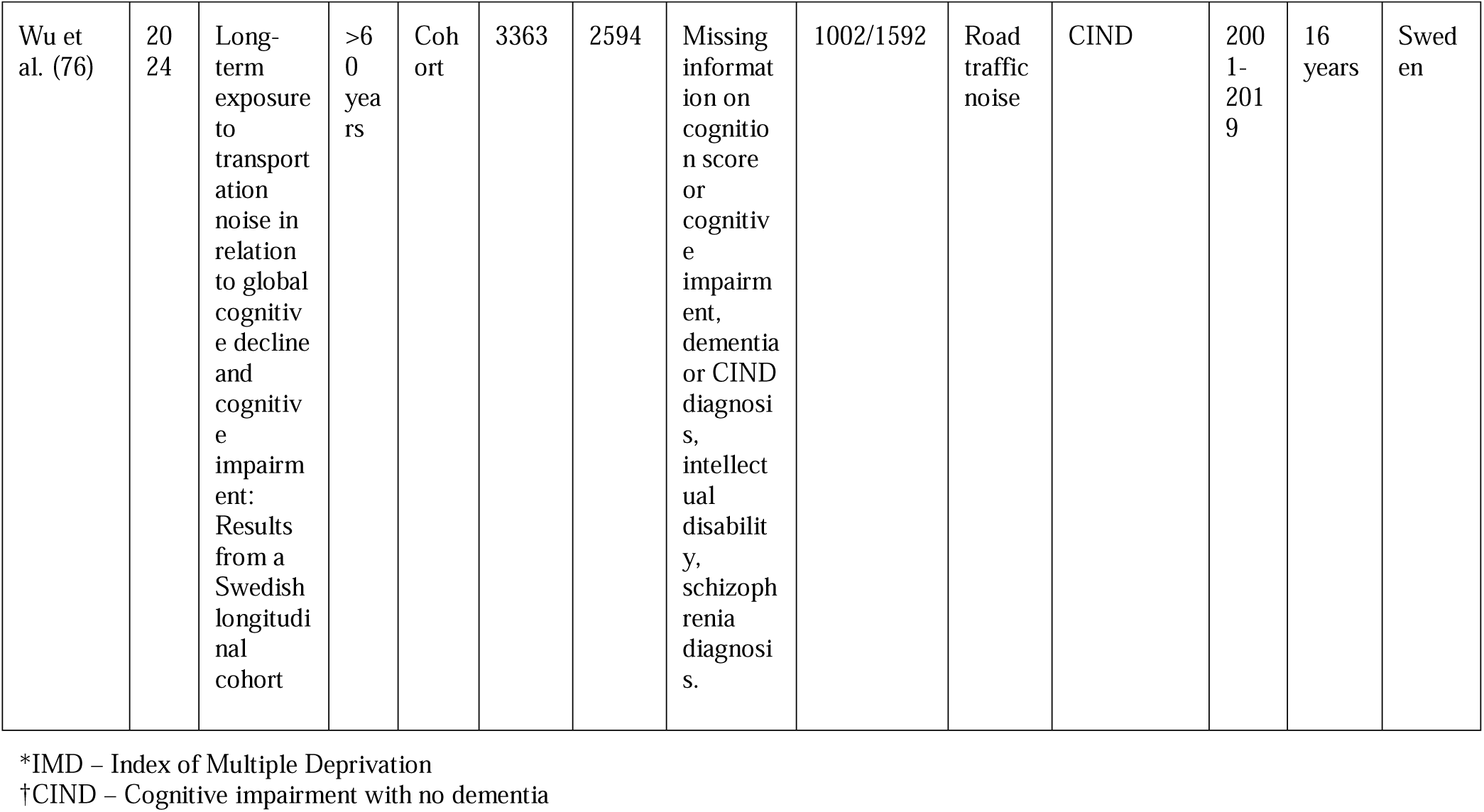
Characteristics of the studies included in the review.

#### 3.2.1 Study design

All the studies were prospective or retrospective cohort studies. Six studies derived their population from larger cohort studies. Cantuaria el al. (50) used the Danish Civil Registration System, Carey et. al (71) used a primary care database (Clinical Practice Research Datalink) in the UK, Wu et al. (76) used the Swedish National study on Aging and Care in Kungsholmen (SNAC-K), and Havyarimana et al. used the UK Biobank. Tuffier et al. (75) used data from the Danish Nurse cohort, and the sample only included female nurses. The average follow-up durations were between 6.5 -16 years for the included studies. The US study (73) had an average follow-up duration of 6.5 years and conducted follow-ups every 12-15 months, while the study with longest follow-up duration had an average follow-up of 16 years with participants assessed every 3-6 years (76). One study (70) contained information of follow-ups conducted every 5 years during the study period, while two others (50, 71) used information from yearly follow-ups.

#### 3.2.2 Exposure

All eight included studies estimated residential road traffic noise exposure using computer-based modelling linked to residential addresses. The models were generated using validated methods and established noise mapping software, in accordance with national or regional noise mapping guidelines. Noise exposure metrics varied with studies reporting equivalent continuous sound levels (Leq or LAeq 24h), day-evening-night levels (Lden), or night-time levels (Lnight).

Leq/LAeq represents the equivalent continuous sound pressure level over a defined time period, A-weighted to better reflect human hearing. Lden is a 24 hour average that adds penalties for noise during the evening and night (77), while Lnight specifically captures noise between 23:00 and 07:00, a period when participants are most likely to be at home and potentially vulnerable to noise-related sleep disturbance (71). Exposure classification approaches varied. Six studies (50, 70, 71, 73, 74, 76) categorised noise exposure, typically defining “high” exposure as ≥ 50–55 dB (Leq, Lden, or Lnight) and “low” exposure as < 50 dB. Category cut-offs were largely consistent across studies, although specific thresholds varied slightly depending on the noise metric used. Two of these studies (74, 76) also analysed noise exposure as a continuous variable, enabling estimation of risk per 10 dB increment. Two studies (72, 75) only analysed noise exposure as a continuous variable.

#### 3.2.3 Outcome

Six studies (50, 70, 71, 73–75) examined dementia incidence as their primary outcome. One study (72) assessed cognitive decline using global cognitive scores, and another study (76) assessed CIND. Ascertainment via routine health/administrative records

Four studies (50, 71, 74, 75) identified dementia from linked clinical and administrative data. This offered complete national coverage and consistent coding over long follow-up, supporting large case counts and narrow CIs. However, registry algorithms may under-ascertain early or atypical cases and offer limited clinical detail compared to follow-up clinical evaluations. The remaining four studies (70, 72, 73, 76) established outcomes through repeated cognitive testing and clinical evaluations.

These studies provide clinically rich, adjudicated outcomes and can capture earlier cognitive changes. However, the resource-intensive process limited the sample sizes and resulted in wider CIs.

The diagnosis of dementia is a clinical decision made based on internationally accepted criteria that include substantial cognitive decline in one or more cognitive domains along with interference in performing daily activities. Cognitive decline is a reduction in cognitive capabilities, often assessed using a cognitive score, and constitutes a major criterion in dementia. The outcomes were diagnosed using standard, internationally accepted diagnostic tools. Two studies (70, 73) used DSM-4 (65) and DSM-5 (20) criteria while four others (50, 71, 74, 75) used ICD-10 (66) criteria. The remaining studies (72, 76) derived standardised global cognitive scores from neuropsychiatric tests and compared intra-individually at baseline and follow-up to assess decline in cognitive scores.

Alzheimer’s disease and vascular dementia were the most commonly reported subtypes when examined separately. Detailed outcome assessment information is provided in Table 4.

#### 3.2.4 Confounders

The confounding factors identified by each study varied slightly. Age, physical activity, smoking, alcohol, air pollution, BMI, education, and certain comorbidities such as hypertension, diabetes and metabolic syndrome were identified by all studies.

#### 3.2.5 Context

Seven studies were conducted within Europe (two each in Sweden, Denmark and the UK, and one in Germany), and one was from the United States of America. The research conducted in USA studied a population of Mexican-Americans in the country (73).

#### 3.2.6 Publication date

All the studies were published between 2018-2025 but used data collected before 2021.

### 3.3 Quality Assessment

The methodological quality of the studies was assessed using the ROBINS-E tool. Studies were assessed over seven domains and graded for overall risk of bias as low, some concerns, high and very high risk as per the tool (68). Six studies were deemed to pose ‘some concerns’ as there was a lack of clarity regarding the accuracy of results due to the presence of missing outcome, exposure, or covariate data. These studies used a complete case analysis to account for missing data. There was insufficient evidence available from the studies to determine whether the results were not affected by the incomplete data. One study (72) was deemed to be ‘high’ risk due to missing data in exposure, outcome and covariates, as well as concerns regarding outcome measurement. One study (50) was judged to have a ‘low’ risk of bias in all domains. The results for each domain and reasons for classifications are reported in Table 3.

**Table 3:** Risk of bias summary for each study with justification for overall rating using the ROBINS-E tool. (**68**)

| Study | Overall Risk of Bias | Domains with risk of bias* | Reasons |
| --- | --- | --- | --- |
| Andersson et al. | Some concerns | D1 | Some important confounders like |
| (2018) (70) |  |  | residence area, occupation, ethnicity and socioeconomic were not controlled for. |
| Cantuaria et al. (2021) (50) | Low risk | None | None. |
| Carey et al. (2018) (71) | Some concerns | D5 | 6% of participants were excluded from the analysis due to missing exposure data. |
| Ogurtsova et al. (2023) (72) | High risk | D5 | Missing exposure, outcome and covariate data that may have affected results despite a complete case analysis. |
| Yu et al. (2020) (73) | Some concerns | D5 | Missing outcome data for 57 participants who were excluded from the analysis. |
| Havyarimana et al. (2025) (74) | Some concerns | D5 | Missing exposure, outcome and covariate data that may have affected results despite a complete case analysis. |
| Tuffier et al. (2024) | Some concerns | D1, D5 | Some confounders not adjusted, and missing |
| (75) |  |  | data in outcome. |
| Wu et al. (2024) (76) | Some concerns | D5 | Missing exposure, outcome and covariate data that may have affected results despite a complete case analysis. |

\*The ROBINS-E tool consists of seven domains that gauge the risk of bias in different areas.
D1 – bias due to confounding
D2 – bias arising from measurement of the exposure
D3 – bias in selection of participants into the study (or into the analysis)
D4 – bias due to post-exposure interventions
D5 – bias due to missing data
D6 – bias arising from measurement of the outcome
D7 – bias in selection of the reported result

**Table 4:**
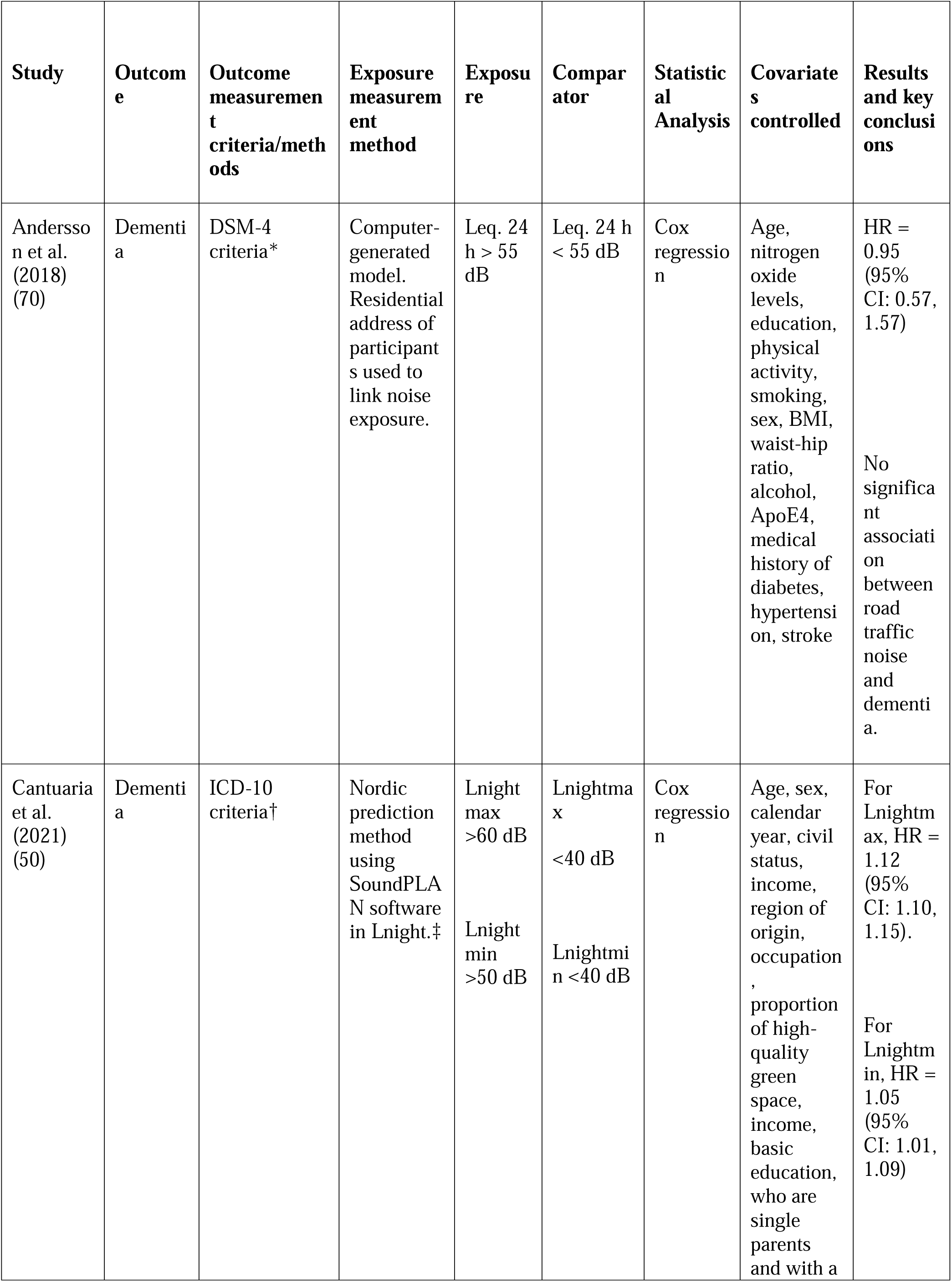
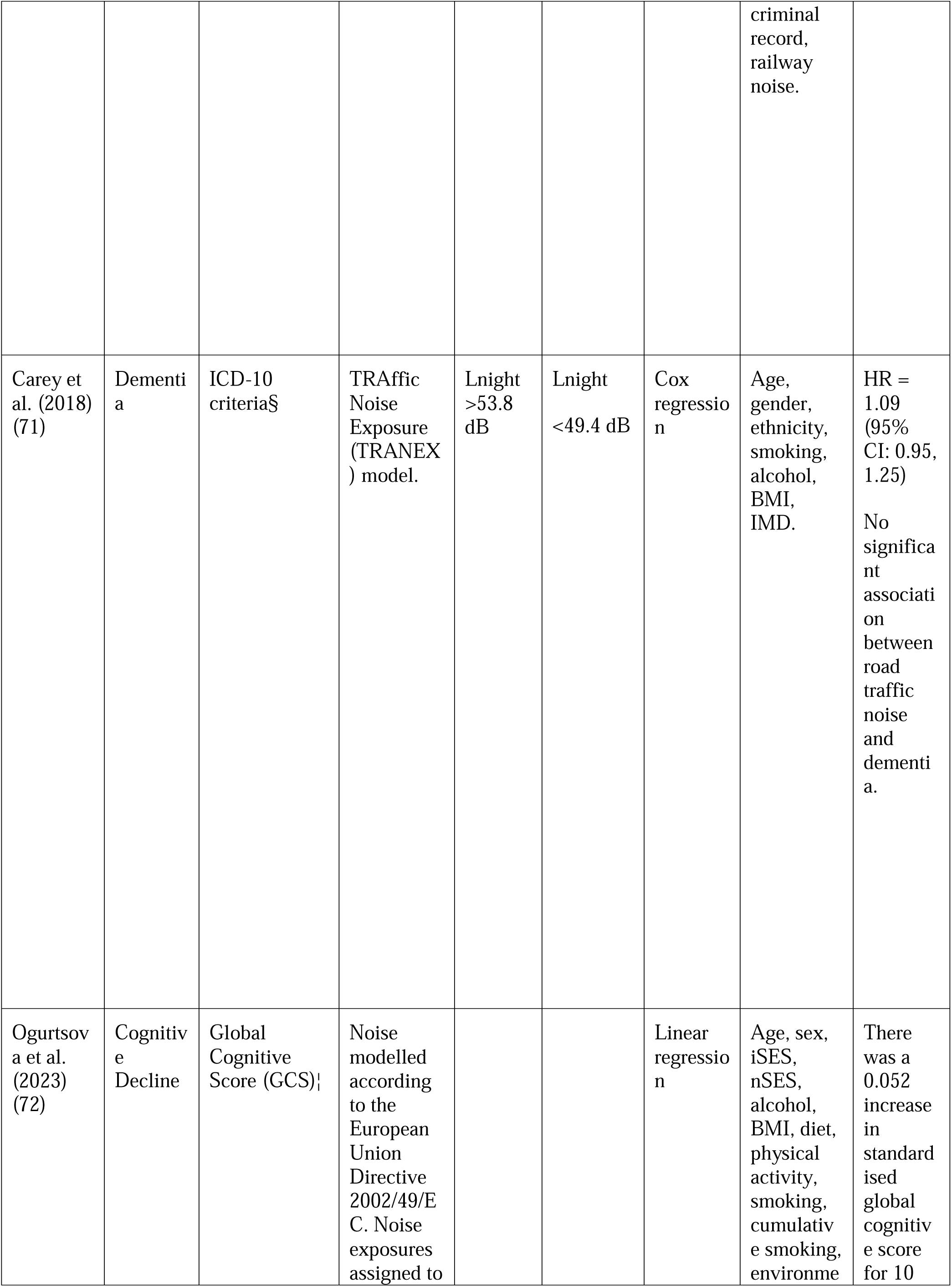
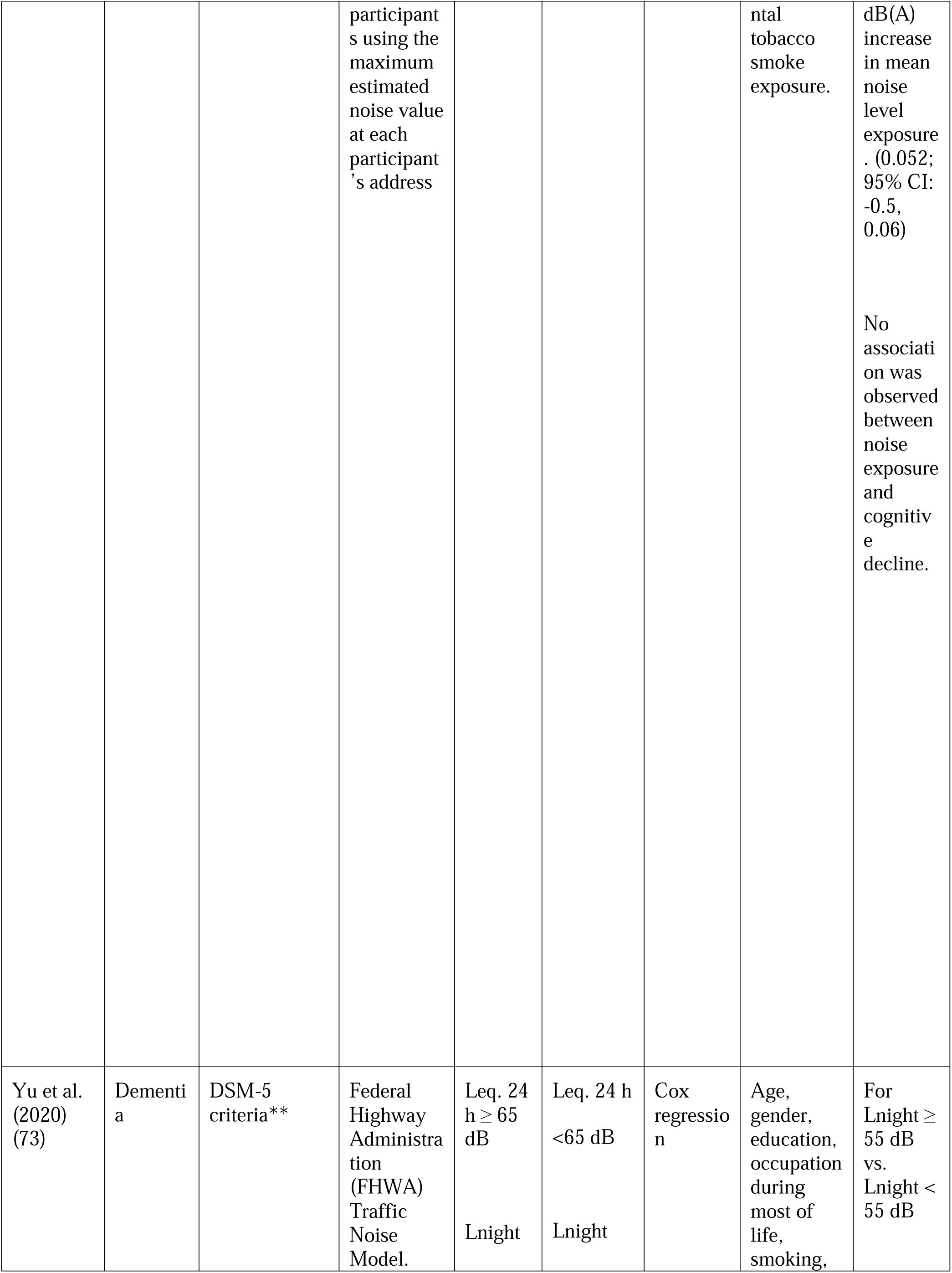
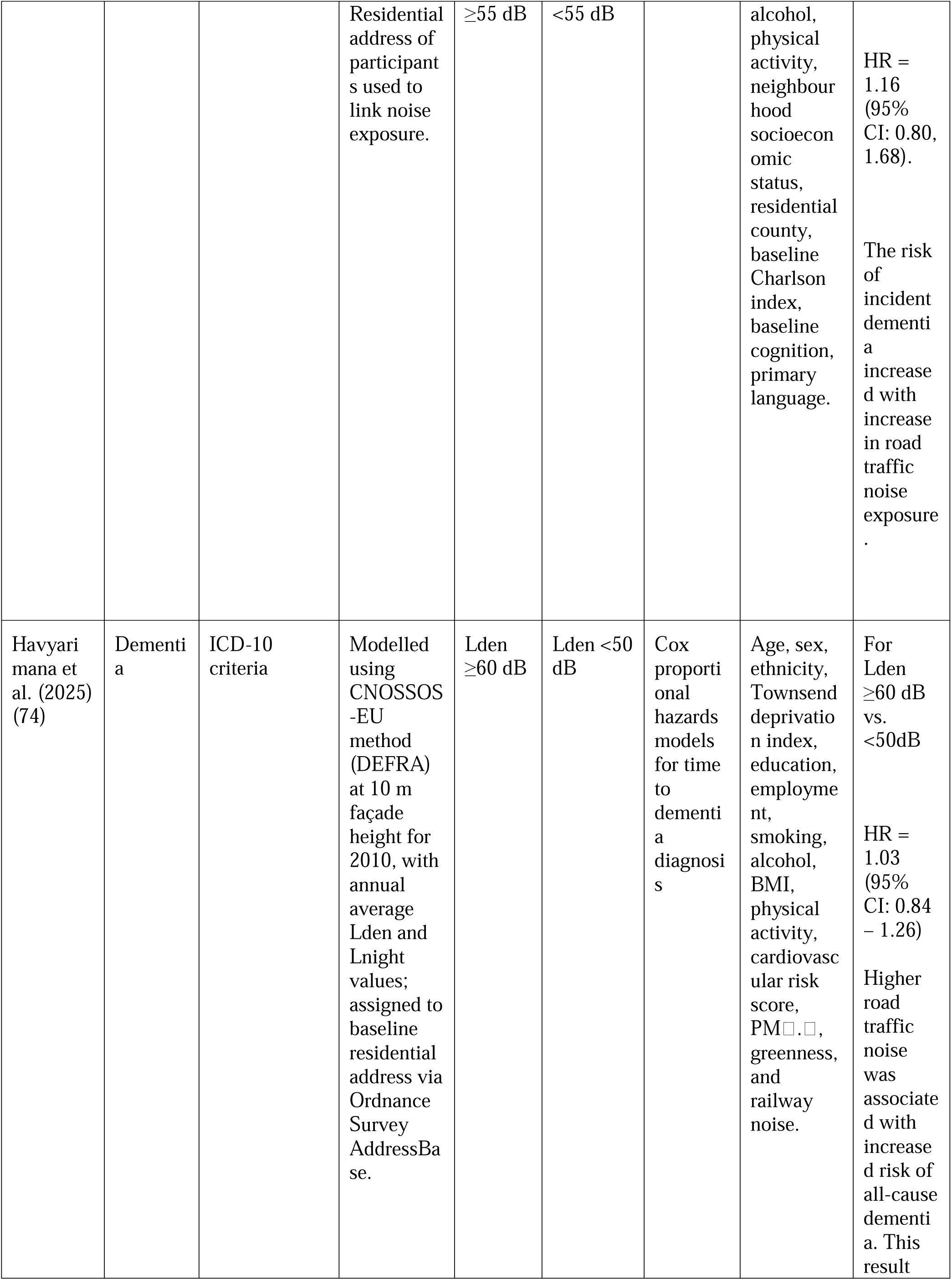
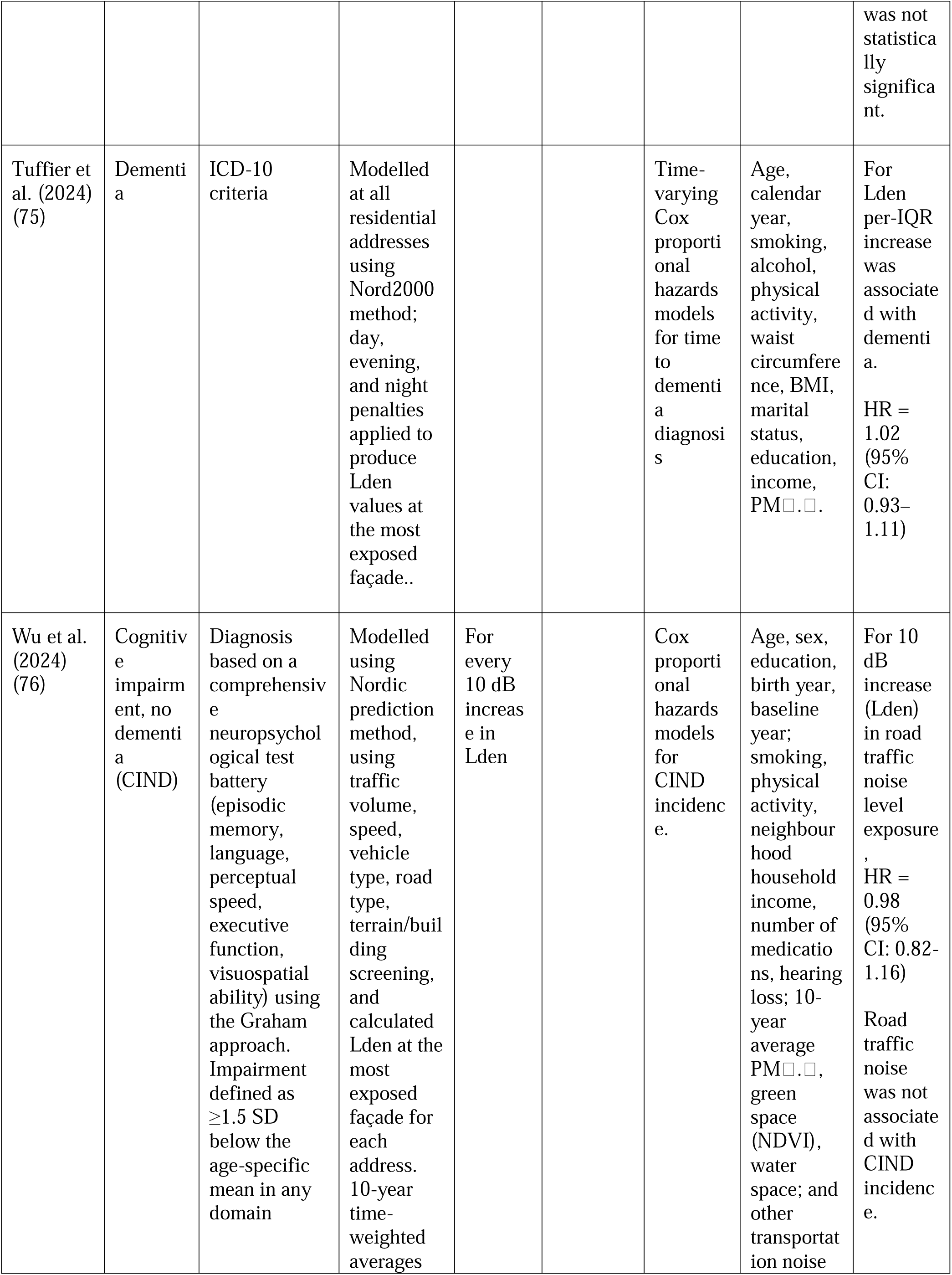
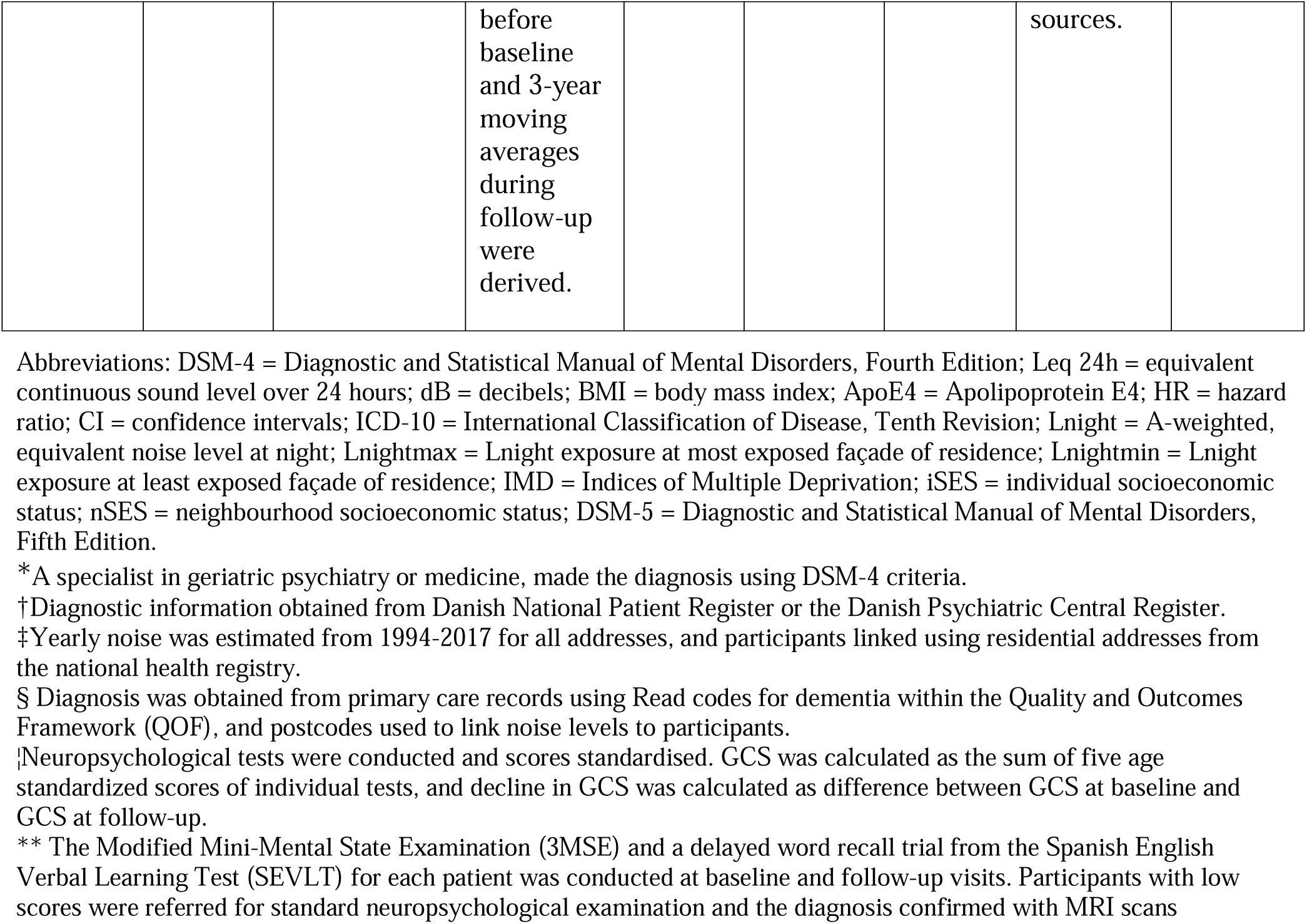
Included studies with outcome, exposure, covariate information, type of statistical analysis and key conclusions.

It was noted that all the studies had large sample sizes, did not use subjective noise measurement tools such as self-reported questionnaires or non-validated surveys, and offered clear diagnostic criteria for outcome assessment. The studies provided adequate information regarding participant characteristics, demographics, methods of exposure and outcome assessment, and statistical analysis. All the studies performed multi-variate analyses using a combination of different confounders that were identified as important. The studies were observed to have low to moderate risk of bias, with residual confounding and bias due to missing data being the only areas of concern. All studies adjusted for age, physical activity, and socioeconomic status. None adjusted for APOE ε4 genotype, and only Wu et al. adjusted for baseline hearing impairment.

### 3.4 Narrative Synthesis

A narrative synthesis was completed to explore the themes and results across the eight included studies. Their results and key conclusions were tabulated and compared in Table 4 to support inferences. All studies modelled noise exposure at the residential address, most often as Lden or Lnight, with cut-offs around 50–55 dB to define higher exposure. Five studies examined dementia incidence as a primary outcome (50, 70, 71, 74, 75), two examined cognitive impairment not meeting dementia criteria or cognitive decline (72, 76), and one examined both dementia and cognitive impairment in a specific population (73).

### Study specific strengths and weaknesses

The included studies varied in design features, exposure assessment, and outcome ascertainment, each contributing unique strengths and limitations. Andersson et al. (2018) conducted a clinical cohort study with dementia diagnoses confirmed by specialists and an average follow-up of seven years, ensuring near-complete capture of incident cases and minimal attrition. However, noise exposure was modelled only at baseline residential addresses, introducing potential misclassification due to unaccounted residential mobility. Cantuaría et al. (2021) leveraged a nationwide Danish cohort with registry-based dementia ascertainment and rich area-level covariates, producing highly precise hazard ratios and strong generalisability. In contrast, Carey et al. (2018) relied on primary care and mortality registry codes, which may have led to under-recording of dementia diagnoses. Ogurtsova et al. (2023) examined a single regional cohort using repeated neuropsychological testing, enhancing validity for cognitive outcomes but facing limited follow-up and possible selection bias. Yu et al. (2020) applied detailed noise modelling linked to geocoded addresses but was constrained by a small sample size, resulting in wide confidence intervals. Havyarimana et al. (2025) analysed the UK Biobank cohort, offering large sample size and long follow-up of approximately 11 years with comprehensive dementia ascertainment, though exposure was modelled only at baseline, again risking misclassification. Tuffier et al. (2024) provided a unique perspective through a nationwide female nurse cohort with detailed residential histories and time-varying exposure estimates, but generalisability was limited by its single-sex occupational sample, and recent exposures were imputed, potentially underestimating variability. Finally, Wu et al. (2024) combined validated neuropsychological assessments over 16 years with detailed noise modelling and extensive covariate adjustment, though its smaller sample size, infrequent follow-up intervals, and lack of dietary or genetic data were notable limitations.

When restricted to dementia incidence, several studies reported positive associations (50, 71, 74). Even where overall estimates for all-cause dementia were close to null, the direction of effect was generally positive. Stronger associations tended to emerge for night-time noise (Lnight) and for metrics based on the least-exposed façade, which may better capture exposure relevant to sleep disruption. In studies examining cognitive decline or CIND, associations were weaker for road traffic noise but more evident for other transportation sources such as rail or aircraft noise. Across cohorts that modelled both air pollution and noise, noise effects often attenuated after pollutant adjustment, yet positive signals persisted in certain subgroups and exposure definitions, suggesting an independent contribution of noise remains plausible.

Several studies did not find statistically significant associations for all-cause dementia, whereas others reported modest positive associations under specific conditions such as Alzheimer’s disease subtype or night-time exposure. Larger cohorts (50, 74) generally produced narrower confidence intervals and more consistent risk estimates, while smaller studies yielded imprecise estimates.

Differences in confounder adjustment were also evident: all studies controlled for key sociodemographic and lifestyle factors, and most included at least one air pollutant. However, psychosocial factors such as social isolation and mental health were unmeasured, and only Wu et al. (76) adjusted for baseline hearing impairment. Two studies (70, 73) did not adjust for air pollution. To avoid overestimating noise effects, this synthesis prioritised fully adjusted hazard ratios, including air pollution where available, and harmonised exposure contrasts (low approximately 50 dB, high 55 dB or greater) across cohorts.

## 4. Discussion

The findings from this review indicate that higher exposure to road traffic noise is associated with a modest but measurable increase in dementia risk compared to lower exposure levels. Across eight longitudinal cohorts involving more than 2.5 million participants, the overall direction of association was positive, particularly when exposure was assessed using night-time noise levels or least-exposed façade metrics, and in large registry-based cohorts. Although effect sizes were small, they are meaningful from a public health perspective because even minor relative risks can translate into substantial population burdens given the ubiquity of traffic noise. This highlights the importance of policy interventions: the World Health Organization recommends that road traffic noise should not exceed 53 dB Lden and 45 dB Lnight to prevent adverse health outcomes, yet these thresholds are frequently exceeded in urban environments. Urban planning measures such as quiet façades, improved building insulation, low-noise road surfaces, and traffic-calming strategies could help mitigate exposure and reduce dementia risk, while also delivering co-benefits for cardiovascular and mental health. Importantly, in studies that adjusted for both noise and air pollution, the association between noise and dementia often attenuated but did not disappear after PM . adjustment (50, 75), suggesting both shared and independent pathways. The nationwide Danish study with façade-specific mapping and adjustment for green space and railway noise provides robust evidence, complemented by the large UK Biobank analysis. Smaller cohorts using repeated clinical assessments tended to yield less precise estimates, but their findings were broadly consistent with the larger studies. These results align with previous research (78, 79). Unlike earlier reviews that included cross-sectional designs or mixed noise sources (47, 49), this review focused exclusively on longitudinal studies, strengthening causal interpretation. Although a meta-analysis was planned, heterogeneity in exposure metrics, outcome definitions, and covariate adjustment precluded meaningful pooling. Despite these challenges, this review offers important insights into the relationship between road traffic noise and dementia, reinforcing the need for integrated environmental health policies.

Chronic exposure to road traffic noise activates the HPA axis, leading to stress hormone dysregulation, sleep disturbance, neuroinflammation, and impaired clearance of neurotoxic proteins. These mechanisms offer a biologically plausible route by which long-term traffic noise contributes to neurodegeneration. Dementia is a progressive disorder with an uncertain duration between noise exposure and onset. Nevertheless, research on other risk factors with similar mechanisms offers an estimate. Studies on known risk factors for dementia, such as hypertension and high cholesterol, suggest that dementia can manifest about 10 years after exposure (80–83). Therefore, long-term studies with regular and complete follow-up assessments provide high-quality evidence. The studies in this review had an average follow-up duration of 6.5 to 16 years, which provide good evidence of association.

All studies in this review used computer-generated models to estimate noise exposure. While direct measurements can also be used, computer-enabled modelling is preferred for long-term monitoring and research studies. Sound level meters and noise dosimeters are validated devices that provide accurate measurements (38), but they are costly and limited in their geographical application. In contrast, modelled noise estimations can efficiently estimate exposure levels over large areas for extended periods (84). However, these models rely on assumptions and can be inaccurate in capturing local variations. Similarly, dementia outcomes were ascertained through national health registries (ICD codes) or electronic health records, which may lack sensitivity for early-stage disease. Standardising diagnostic protocols and integrating cognitive testing batteries could improve comparability across future investigations.

The relationship between road traffic noise and dementia is complicated by the presence of confounders, such as exposure to air pollution, which has not been adequately studied. While some studies have adjusted for air pollution, there is little research on its actual impact on the relationship between noise exposure and dementia. Additionally, studies investigating the effects of air pollution and noise pollution independently are methodologically challenging due to their tendency to occur in combination, and it is unclear whether their combined risks are linear. Certain confounders such as social isolation and psychosocial factors were not adjusted for in any of the studies. Social isolation is linked to age (85) and hearing impairment (86), which are both associated with dementia independently, and may be part of a chain of causation leading to poor neurocognitive outcomes. A study published in 2023 found that social isolation increases the risk of dementia by 30% (59), and this risk was not modified by genetic factors (87). This suggests that other factors, such as psychosocial factors, may also play a role in the development of dementia.

Differences in noise metrics, facade selection (most-vs least-exposed), and exposure time windows likely contribute to varied results. Outcome definitions also varied. Registry-based dementia captures later disease stages with high specificity but lower sensitivity, and cognitive decline and CIND may detect earlier changes but are not equivalent to dementia incidence.

### Strengths of the review

This review critically assessed high-quality cohort studies examining the connection between exposure to road traffic noise and dementia. Following best practices in systematic review methodology, the review incorporated PRISMA guidelines and involved two researchers in the screening, data extraction, and risk of bias assessments. The review only included longitudinal studies, which are the most robust study design to explain this relationship, as randomised controlled trials would be highly unethical and inappropriate in this context. Previous reviews that included other study designs, such as cross-sectional studies, suffered from recall and selection bias, affecting the accuracy of their results.

### Limitations of the review

While the search was extensive, only a few primary studies were available, which limited the number of studies included in the review and increased the heterogeneity between them.

Upon analysing the primary studies included in this review, it was observed that the studies assessed road noise exposure using modelled data, linking predicted noise levels in each area to the participants’ addresses. While cost-effective, this method is prone to misclassification bias as exposure may vary based on factors such as time spent at home, use of personal noise-protection equipment, and sound-proofing in construction. The exposure is usually modelled based on the loudest façade, and this may also result in an overestimation of the exposure (88). Validation sub-studies using personal noise dosimeters are needed to quantify measurement error and refine dose– response estimates. Additionally, all except three studies (50, 75, 76) failed to collect complete residential history, which is an important essential determinant of exposure duration. Four studies (50, 71, 74, 75) used primary care records and health registers, respectively, to obtain dementia diagnosis, which could have been prone to outcome misclassification. Underreporting and underdiagnosis of dementia is also an acute issue in primary care (89).

Exposure metric variability (24-hour Leq versus Lnight or Lden) limited direct comparability and precluded unified dose-response analyses. Leq captures overall daily exposure but may underrepresent nighttime peaks critical for sleep disturbance, whereas Lnight specifically addresses nocturnal exposure at the expense of missing daytime noise burdens. Lden applies a weighted 24-hour average with added penalties for evening and night-time noise to reflect increased sensitivity during these periods, but this aggregation can mask short-term peaks, rely on arbitrary penalty values, and reduce temporal specificity, potentially obscuring the distinct health impacts of daytime versus nocturnal exposure.

Six of the eight studies (71–76) had missing data, and all of them used a complete case analysis. This approach reduces the sample size and may lead to biased results because participants with missing data may differ from those with complete data. A better solution is to impute missing data, which would preserve data from participants with missing information and provide a less biased estimate. Complete case analysis remains a simpler method and can be reasonable when the proportion of missing data is small, as was the case in some cohorts. However, in larger datasets with complex exposure and outcome measures, imputation could improve precision and validity of effect estimates. Two studies (50, 70) reported the same number of participants at baseline and in their main analyses, suggesting minimal or no missingness for variables used in the models. The possibility of bias due to unaccounted confounders is also a concern. None of the studies except Wu et al. (76) adjusted for baseline hearing impairment, a potential confounder that could bias associations if hearing loss both increases susceptibility to noise-induced stress and correlates with cognitive decline. Lastly, the results may not be applicable globally as the studies included were conducted in Europe or North America and only represent a small portion of the world’s population.

### Implications and further research

An important caveat is the scarcity of robust longitudinal investigations examining traffic noise and neurodegeneration. Most existing studies employ cross sectional designs, which preclude causal inference and may be confounded by unmeasured factors. Further high-quality research is needed to explore the impact of road noise on cognitive health outcomes. It is suggested that symptoms of dementia occur long before diagnosis is made (90), and most large cohort studies with cognitive impairment or dementia as outcomes have insufficient follow-up periods for cognitive screening tests. Only a handful of cohort studies have followed participants over extended periods with repeated noise assessments and clinical endpoints, limiting our ability to characterise temporal relationships and dose–response dynamics. Future research should prioritize prospective designs with standardised exposure metrics and long term follow up to more definitively establish a link between chronic traffic noise and dementia development.

All included studies were conducted in high-income countries, predominantly in Europe and North America. Urban form, vehicle fleet composition, and noise modelling approaches differ substantially in low- and middle-income settings. Thus, the urgent need for prospective studies in Asia, Africa, and Latin America, where traffic mixtures and socioeconomic factors may modify exposure– response relationships. Pollution exposure is influenced by geography and is not consistent across all regions. Ethnicity is linked with an increased risk of dementia (91), and some ethnic groups such as African-Americans and Caribbean-Hispanics may be more susceptible to developing dementia than others (92, 93). Urbanisation and population growth have led to an expansion of road networks and increase in traffic (94), which exposes people living in densely populated areas and those without access to developed public transport systems to higher levels of road traffic noise (95). Hence, low socioeconomic status is also associated with an increased exposure to traffic noise (96, 97). Vehicle honking, which adds 2 to 5 dB(A) to noise levels, is a significant contributor to road traffic noise (98, 99) and can be influenced by cultural differences where honking is used as a form of non-verbal communication (100).

Although traffic noise has been linked to cognitive decline, none of the cohort studies in our review adjusted for the APOE ε4 allele, the most significant genetic risk factor for dementia (101, 102).

Future cohorts should collect biospecimens for genotyping and incorporate stratified analyses or interaction testing to determine whether noise-related dementia risk is modified by APOE status. Future studies must also harmonise noise metrics (e.g., prioritize Leq24h or Lden) and include audiometric assessments to control for hearing status. Dementia sub-types must be disaggregated, and consistent modelling strategies must be used to clarify independent effects of covariates.

## 5. Conclusion

In conclusion, exposure to high levels of road traffic noise is associated with an increase in the risk of dementia or cognitive decline based on existing evidence. While heterogeneity precluded a defensible meta-estimate, convergent patterns across designs indicate that noise levels in the common policy range (≈50–55 dB and above) may be relevant for dementia risk especially where night-time exposure is involved. Future research should standardise noise exposure metrics, integrate APOE ε4 genotyping and hearing impairment adjustments, validate modeled exposures with personal dosimetry, and expand prospective cohorts to low- and middle-income settings.

From a public health perspective, interventions to reduce night-time transportation noise, through the installation of quiet façades, better insulation, and urban planning, are justified given potential neurological, cardiovascular, and sleep benefits. This may have far-reaching implications on the planning of transport infrastructure and development of policies to mitigate or prevent adverse health outcomes due to exposure to high levels of road traffic noise.

## 6. Author Contributions

E.S. and N.D. conceived the study and designed the search strategy. E.S. and N.D. conducted the literature search, and conducted title and abstract, and full text screening. Data extraction was carried out by E.S. and verified by N.D. Both authors (E.S. and N.D.) analyzed the data and interpreted the findings. E.S and N.D. wrote the manuscript and approved the final version.

## Data Availability

All data produced in the present study are available upon reasonable request to the authors.

## Acknowledgements

We thank Dr. Suzanne Bartington for her suggestions and guidance in course of this research. We also thank the staff at the Department of Applied Health Research, University of Birmingham for their valuable support.

## References

1. Liu F, Jiang S, Kang J, Wu Y, Yang D, Meng Q, et al. On the definition of noise. Humanities and Social Sciences Communications. 2022;9(1):406.

2. Khademi G, Imani B. Noise pollution in intensive care units: a systematic review article. Reviews in Clinical Medicine. 2015;2(2):58–64.

3. Lee Y, Lee S, Lee W. Occupational and Environmental Noise Exposure and Extra-Auditory Effects on Humans: A Systematic Literature Review. Geohealth. 2023;7(6).

4. Muralikrishna IV, Manickam V. Chapter Eleven - Principles and Design of Water Treatment. In: Muralikrishna IV, Manickam V, editors. Environmental Management: Butterworth-Heinemann; 2017. p. 209–48.

5. Welch D, Shepherd D, Dirks KN, Reddy R. Health effects of transport noise. Transport Reviews. 2023;43(6):1190–210.

6. Basner M, Babisch W, Davis A, Brink M, Clark C, Janssen S, et al. Auditory and non-auditory effects of noise on health. Lancet. 2014;383(9925):1325-32.

7. Oguntunde PE, Okagbue HI, Oguntunde OA, Odetunmibi OO. A Study of Noise Pollution Measurements and Possible Effects on Public Health in Ota Metropolis, Nigeria. Open Access Maced J Med Sci. 2019;7(8):1391–5.

8. Hinchcliffe R. Review: Global perspective of noise-induced hearing loss as exemplified by Nigeria. Journal of Audiological Medicine. 2002;11:1–24.

9. Nichols E, Steinmetz JD, Vollset SE, Fukutaki K, Chalek J, Abd-Allah F, et al. Estimation of the global prevalence of dementia in 2019 and forecasted prevalence in 2050: an analysis for the Global Burden of Disease Study 2019. The Lancet Public Health. 2022;7(2):e105–e25.

10. Prince M, Guerchet M, Prina M. The Epidemiology and Impact of Dementia - Current State and Future Trends. WHO Thematic Briefing2015.

11. World Health Organization, Brown L. Burden of Disease from Environmental Noise. Quantification of Healthy Life Years Lost in Europe. 2011.

12. Department for Infrastructure and Transport - South Australia. Road Traffic Noise Guidelines - EHTM Attachment 7A. 2001.

13. Selander J, Bluhm G, Theorell T, Pershagen G, Babisch W, Seiffert I, et al. Saliva cortisol and exposure to aircraft noise in six European countries. Environ Health Perspect. 2009;117(11):1713–7.

14. Münzel T, Gori T, Babisch W, Basner M. Cardiovascular effects of environmental noise exposure. Eur Heart J. 2014;35(13):829–36.

15. Liu X-q, Huang J, Song C, Zhang T-l, Liu Y-p, Yu L. Neurodevelopmental toxicity induced by PM2.5 Exposure and its possible role in Neurodegenerative and mental disorders. Human & Experimental Toxicology. 2023;42:09603271231191436.

16. Boyacioglu N, Ozkan S. The Effect of Noise in the Intensive Care Unit on the Oxidative Stress Response in Rats. Biological Research For Nursing. 2020;22(3):397–402.

17. Cheng L, Wang S-H, Chen Q-C, Liao X-M. Moderate noise induced cognition impairment of mice and its underlying mechanisms. Physiology & Behavior. 2011;104(5):981–8.

18. Cui B, Zhu L, She X, Wu M, Ma Q, Wang T, et al. Chronic noise exposure causes persistence of tau hyperphosphorylation and formation of NFT tau in the rat hippocampus and prefrontal cortex. Experimental neurology. 2012;238(2):122–9.

19. Basner M, Müller U, Elmenhorst EM. Single and combined effects of air, road, and rail traffic noise on sleep and recuperation. Sleep. 2011;34(1):11–23.

20. American Psychiatric Association. Diagnostic and Statistical Manual of Mental Disorders, Fifth Edition (DSM-5). Arlington, VA: American Psychiatric Association Publishing; 2013.

21. Mark RE, Brehmer Y. Preclinical Alzheimer’s dementia: a useful concept or another dead end? European Journal of Ageing. 2022;19(4):997–1004.

22. Amieva H, Le Goff M, Millet X, Orgogozo JM, Pérès K, Barberger-Gateau P, et al. Prodromal Alzheimer’s disease: successive emergence of the clinical symptoms. Ann Neurol. 2008;64(5):492–8.

23. Warren A. Behavioral and Psychological Symptoms of Dementia as a Means of Communication: Considerations for Reducing Stigma and Promoting Person-Centered Care. Frontiers in Psychology. 2022;13.

24. Flier WMvd, Scheltens P. Epidemiology and risk factors of dementia. Journal of Neurology, Neurosurgery & Psychiatry. 2005;76(suppl 5):v2.

25. Bartolone SN, Sharma P, Chancellor MB, Lamb LE. Urinary Incontinence and Alzheimer’s Disease: Insights From Patients and Preclinical Models. Front Aging Neurosci. 2021;13:777819.

26. Berr C, Wancata J, Ritchie K. Prevalence of dementia in the elderly in Europe. European Neuropsychopharmacology. 2005;15(4):463–71.

27. Burks HB, des Bordes JKA, Chadha R, Holmes HM, Rianon NJ. Quality of Life Assessment in Older Adults with Dementia: A Systematic Review. Dementia and Geriatric Cognitive Disorders. 2021;50(2):103–10.

28. Ritchie K, Lovestone S. The dementias. The Lancet. 2002;360(9347):1759–66.

29. Deary IJ, Corley J, Gow AJ, Harris SE, Houlihan LM, Marioni RE, et al. Age-associated cognitive decline. Br Med Bull. 2009;92:135–52.

30. Hay SI, Abajobir AA, Abate KH, Abbafati C, Abbas KM, Abd-Allah F, et al. Global, regional, and national disability-adjusted life-years (DALYs) for 333 diseases and injuries and healthy life expectancy (HALE) for 195 countries and territories, 1990&#x2013;2016: a systematic analysis for the Global Burden of Disease Study 2016. The Lancet. 2017;390(10100):1260–344.

31. Dominguez LJ, Veronese N, Vernuccio L, Catanese G, Inzerillo F, Salemi G, et al. Nutrition, Physical Activity, and Other Lifestyle Factors in the Prevention of Cognitive Decline and Dementia. Nutrients. 2021;13(11).

32. Baumgart M, Snyder HM, Carrillo MC, Fazio S, Kim H, Johns H. Summary of the evidence on modifiable risk factors for cognitive decline and dementia: A population-based perspective. Alzheimers Dement. 2015;11(6):718–26.

33. Arvanitakis Z, Shah RC, Bennett DA. Diagnosis and Management of Dementia: Review. Jama. 2019;322(16):1589–99.

34. Garre-Olmo J, Ponjoan A, Inoriza JM, Blanch J, Sánchez-Pérez I, Cubí R, et al. Survival, effect measures, and impact numbers after dementia diagnosis: a matched cohort study. Clin Epidemiol. 2019;11:525–42.

35. Wimo A, Seeher K, Cataldi R, Cyhlarova E, Dielemann JL, Frisell O, et al. The worldwide costs of dementia in 2019. Alzheimer’s & Dementia. 2023;19(7):2865–73.

36. Jephcote C, Clark SN, Hansell AL, Jones N, Chen Y, Blackmore C, et al. Spatial assessment of the attributable burden of disease due to transportation noise in England. Environment International. 2023;178:107966.

37. Jarosińska D, Héroux M, Wilkhu P, Creswick J, Verbeek J, Wothge J, et al. Development of the WHO Environmental Noise Guidelines for the European Region: An Introduction. Int J Environ Res Public Health. 2018;15(4).

38. World Health Organization. Environmental Health Criteria 12 - Noise. Environmental Health Criteria 12 [Internet]. 1980:[87 p.]. Available from: https://apps.who.int/iris/bitstream/handle/10665/39458/9241540729-eng.pdf.

39. World Health Organization. Recommendations of the WHO Environmental noise guidelines for the European Region (Table 8.11). WHO Housing and Health Guidelines. Geneva. 2018.

40. Tzivian L, Dlugaj M, Winkler A, Weinmayr G, Hennig F, Fuks KB, et al. Long-Term Air Pollution and Traffic Noise Exposures and Mild Cognitive Impairment in Older Adults: A Cross-Sectional Analysis of the Heinz Nixdorf Recall Study. Environ Health Perspect. 2016;124(9):1361–8.

41. Crous-Bou M, Gascon M, Gispert JD, Cirach M, Sánchez-Benavides G, Falcon C, et al. Impact of urban environmental exposures on cognitive performance and brain structure of healthy individuals at risk for Alzheimer’s dementia. Environ Int. 2020;138:105546.

42. Fuks KB, Wigmann C, Altug H, Schikowski T. Road Traffic Noise at the Residence, Annoyance, and Cognitive Function in Elderly Women. Int J Environ Res Public Health. 2019;16(10).

43. Linares C, Culqui D, Carmona R, Ortiz C, Díaz J. Short-term association between environmental factors and hospital admissions due to dementia in Madrid. Environ Res. 2017;152:214–20.

44. Chen H, Kwong JC, Copes R, Tu K, Villeneuve PJ, van Donkelaar A, et al. Living near major roads and the incidence of dementia, Parkinson’s disease, and multiple sclerosis: a population-based cohort study. Lancet. 2017;389(10070):718–26.

45. Tzivian L, Jokisch M, Winkler A, Weimar C, Hennig F, Sugiri D, et al. Associations of long-term exposure to air pollution and road traffic noise with cognitive function-An analysis of effect measure modification. Environ Int. 2017;103:30–8.

46. Ju YJ, Lee JE, Lee SY. Perceived environmental pollution and subjective cognitive decline (SCD) or SCD-related functional difficulties among the general population. Environ Sci Pollut Res Int. 2021;28(24):31289–300.

47. Clark C, Crumpler C, Notley AH. Evidence for Environmental Noise Effects on Health for the United Kingdom Policy Context: A Systematic Review of the Effects of Environmental Noise on Mental Health, Wellbeing, Quality of Life, Cancer, Dementia, Birth, Reproductive Outcomes, and Cognition. Int J Environ Res Public Health. 2020;17(2).

48. Hegewald J, Schubert M, Freiberg A, Romero Starke K, Augustin F, Riedel-Heller SG, et al. Traffic Noise and Mental Health: A Systematic Review and Meta-Analysis. Int J Environ Res Public Health. 2020;17(17).

49. Meng L, Zhang Y, Zhang S, Jiang F, Sha L, Lan Y, et al. Chronic Noise Exposure and Risk of Dementia: A Systematic Review and Dose-Response Meta-Analysis. Front Public Health. 2022;10:832881.

50. Cantuaria ML, Waldorff FB, Wermuth L, Pedersen ER, Poulsen AH, Thacher JD, et al. Residential exposure to transportation noise in Denmark and incidence of dementia: national cohort study. BMJ. 2021;374:n1954.

51. Fayosse A, Nguyen D-P, Dugravot A, Dumurgier J, Tabak AG, Kivimäki M, et al. Risk prediction models for dementia: role of age and cardiometabolic risk factors. BMC Medicine. 2020;18(1):107.

52. Daviglus ML, Bell CC, Berrettini W, Bowen PE, Connolly ES, Jr., Cox NJ, et al. National Institutes of Health State-of-the-Science Conference statement: preventing alzheimer disease and cognitive decline. Ann Intern Med. 2010;153(3):176–81.

53. Gong J, Harris K, Lipnicki DM, Castro-Costa E, Lima-Costa MF, Diniz BS, et al. Sex differences in dementia risk and risk factors: Individual-participant data analysis using 21 cohorts across six continents from the COSMIC consortium. Alzheimer’s & Dementia. 2023;19(8):3365–78.

54. Letellier N, Yang J-A, Cavaillès C, Casey JA, Carrasco-Escobar G, Zamora S, et al. Aircraft and road traffic noise, insulin resistance, and diabetes: The role of neighborhood socioeconomic status in San Diego County. Environmental Pollution. 2023;335:122277.

55. Dale LM, Goudreau S, Perron S, Ragettli MS, Hatzopoulou M, Smargiassi A. Socioeconomic status and environmental noise exposure in Montreal, Canada. BMC Public Health. 2015;15:205.

56. Petersen JD, Wehberg S, Packness A, Svensson NH, Hyldig N, Raunsgaard S, et al. Association of Socioeconomic Status With Dementia Diagnosis Among Older Adults in Denmark. JAMA Netw Open. 2021;4(5):e2110432.

57. Wu J, Zou C, He S, Sun X, Wang X, Yan Q. Traffic noise exposure of high-rise residential buildings in urban area. Environmental Science and Pollution Research. 2019;26(9):8502–15.

58. Shukla A, Harper M, Pedersen E, Goman A, Suen JJ, Price C, et al. Hearing Loss, Loneliness, and Social Isolation: A Systematic Review. Otolaryngol Head Neck Surg. 2020;162(5):622–33.

59. Huang AR, Roth DL, Cidav T, Chung S-E, Amjad H, Thorpe Jr RJ, et al. Social isolation and 9-year dementia risk in community-dwelling Medicare beneficiaries in the United States. Journal of the American Geriatrics Society. 2023;71(3):765–73.

60. World Health Organization. Burden of disease from environmental noise: quantification of healthy life years lost in Europe. Copenhagen: World Health Organization Regional Office for Europe; 2011.

61. Page MJ, McKenzie JE, Bossuyt PM, Boutron I, Hoffmann TC, Mulrow CD, et al. The PRISMA 2020 statement: an updated guideline for reporting systematic reviews. BMJ. 2021;372:n71.

62. Huang L, Zhang Y, Wang Y, Lan Y. Relationship Between Chronic Noise Exposure, Cognitive Impairment, and Degenerative Dementia: Update on the Experimental and Epidemiological Evidence and Prospects for Further Research. J Alzheimers Dis. 2021;79(4):1409–27.

63. The EndNote Team. EndNote. Philadelphia, PA: Clarivate; 2013.

64. Covidence systematic review software. Veritas Health Innovation. 2023. Available from: www.covidence.org.

65. American Psychiatric Association. Diagnostic and Statistical Manual of Mental Disorders, Fourth Edition. Washington, DC: American Psychiatric Association Publishing; 2000.

66. World Health Organization. International Classification of Diseases Tenth revision ed. Geneva: World Health Organization; 2019.

67. World Health Organization. International Statistical Classification of Diseases and Related Health Problems 10th Revision (ICD 10). Geneva. World Health Organization. 1993.

68. Higgins J, Morgan R, Rooney A, Taylor K, Thayer K, Silva R, et al. Risk Of Bias In Non-randomized Studies - of Exposure (ROBINS-E). 2022 [Available from: https://www.riskofbias.info/welcome/robins-e-tool.

69. Bero L, Chartres N, Diong J, Fabbri A, Ghersi D, Lam J, et al. The risk of bias in observational studies of exposures (ROBINS-E) tool: concerns arising from application to observational studies of exposures. Syst Rev. 2018;7(1):242.

70. Andersson J, Oudin A, Sundström A, Forsberg B, Adolfsson R, Nordin M. Road traffic noise, air pollution, and risk of dementia - results from the Betula project. Environ Res. 2018;166:334–9.

71. Carey IM, Anderson HR, Atkinson RW, Beevers SD, Cook DG, Strachan DP, et al. Are noise and air pollution related to the incidence of dementia? A cohort study in London, England. BMJ Open. 2018;8(9):e022404.

72. Ogurtsova K, Soppa VJ, Weimar C, Jöckel K-H, Jokisch M, Hoffmann B. Association of long-term air pollution and ambient noise with cognitive decline in the Heinz Nixdorf Recall study. Environmental Pollution. 2023;331:121898.

73. Yu Y, Mayeda ER, Paul KC, Lee E, Jerrett M, Su J, et al. Traffic-related Noise Exposure and Late-life Dementia and Cognitive Impairment in Mexican-Americans. Epidemiology. 2020;31(6):771–8.

74. Havyarimana E, Gong X, Jephcote C, Johnson S, Suri S, Xie W, et al. Residential exposure to road and railway traffic noise and incidence of dementia: The UK Biobank cohort study. Environmental Research. 2025;279:121787.

75. Tuffier S, Zhang J, Bergmann M, So R, Napolitano GM, Cole-Hunter T, et al. Long-term exposure to air pollution and road traffic noise and incidence of dementia in the Danish Nurse Cohort. Alzheimer’s & Dementia. 2024;20(6):4080–91.

76. Wu J, Grande G, Pyko A, Laukka EJ, Pershagen G, Ögren M, et al. Long-term exposure to transportation noise in relation to global cognitive decline and cognitive impairment: Results from a Swedish longitudinal cohort. Environment International. 2024;185:108572.

77. European Environment A. Day-evening-night level (Lden). 2025.

78. Paul KC, Haan M, Mayeda ER, Ritz BR. Ambient Air Pollution, Noise, and Late-Life Cognitive Decline and Dementia Risk. Annu Rev Public Health. 2019;40:203–20.

79. Thompson R, Smith RB, Bou Karim Y, Shen C, Drummond K, Teng C, et al. Noise pollution and human cognition: An updated systematic review and meta-analysis of recent evidence. Environment International. 2022;158:106905.

80. Sierra C. Hypertension and the Risk of Dementia. Front Cardiovasc Med. 2020;7:5.

81. Cleveland Clinic. Alzheimer’s Disease. The Cleveland Clinic Health Information Center [Internet] 2022 [cited 2023 August 10]. Available from: https://my.clevelandclinic.org/health/diseases/9164-alzheimers-disease.

82. Kivipelto M, Helkala E-L, Laakso MP, Hänninen T, Hallikainen M, Alhainen K, et al. Midlife vascular risk factors and Alzheimer&#039;s disease in later life: longitudinal, population based study. BMJ. 2001;322(7300):1447.

83. Launer LJ, Ross GW, Petrovitch H, Masaki K, Foley D, White LR, et al. Midlife blood pressure and dementia: the Honolulu-Asia aging study. Neurobiol Aging. 2000;21(1):49–55.

84. Adegoke O, Gbadamosi S, Owolabi I, Nwulu N. Noise Measurement, Characterization, and Modeling for Broadband Indoor Power Communication System: A Comprehensive Survey. Energies. 2023;16.

85. Moormann KI, Pabst A, Bleck F, Löbner M, Kaduszkiewicz H, van der Leeden C, et al. Social isolation in the oldest-old: determinants and the differential role of family and friends. Social Psychiatry and Psychiatric Epidemiology. 2023.

86. Jiang F, Kuper H, Zhou C, Qin W, Xu L. Relationship between hearing loss and depression symptoms among older adults in China: The mediating role of social isolation and loneliness. Int J Geriatr Psychiatry. 2022;37(6).

87. Elovainio M, Lahti J, Pirinen M, Pulkki-Råback L, Malmberg A, Lipsanen J, et al. Association of social isolation, loneliness and genetic risk with incidence of dementia: UK Biobank Cohort Study. BMJ Open. 2022;12(2):e053936.

88. Vienneau D, Héritier H, Foraster M, Eze IC, Schaffner E, Thiesse L, et al. Façades, floors and maps – Influence of exposure measurement error on the association between transportation noise and myocardial infarction. Environment International. 2019;123:399–406.

89. Amjad H, Roth DL, Sheehan OC, Lyketsos CG, Wolff JL, Samus QM. Underdiagnosis of Dementia: an Observational Study of Patterns in Diagnosis and Awareness in US Older Adults. J Gen Intern Med. 2018;33(7):1131–8.

90. Floud S, Balkwill A, Sweetland S, Brown A, Reus EM, Hofman A, et al. Cognitive and social activities and long-term dementia risk: the prospective UK Million Women Study. Lancet Public Health. 2021;6(2):e116–e23.

91. Shiekh SI, Cadogan SL, Lin LY, Mathur R, Smeeth L, Warren-Gash C. Ethnic Differences in Dementia Risk: A Systematic Review and Meta-Analysis. J Alzheimers Dis. 2021;80(1):337–55.

92. Kornblith E, Bahorik A, Boscardin WJ, Xia F, Barnes DE, Yaffe K. Association of Race and Ethnicity With Incidence of Dementia Among Older Adults. JAMA. 2022;327(15):1488–95.

93. Mehta KM, Yeo GW. Systematic review of dementia prevalence and incidence in United States race/ethnic populations. Alzheimers Dement. 2017;13(1):72–83.

94. Dietrich S. Guidelines for Environmental Noise Management in Developing Countries. In: Mia S, editor. Management of Noise Pollution. Rijeka: IntechOpen; 2023.

95. Gilani TA, Mir MS. Association of road traffic noise exposure and prevalence of coronary artery disease: A cross-sectional study in North India. Environmental Science and Pollution Research. 2021;28(38):53458–77.

96. Karandagh ST, Alimohammadi I, Moatar F, Kanrash FA. Association between noise annoyance and socioeconomic status of the employees in an electrical panel manufacturer. Applied Acoustics. 2021;176:107889.

97. Hoffmann B, Robra BP, Swart E. Social inequality and noise pollution by traffic in the living environment--an analysis by the German Federal Health Survey (Bundesgesundheitssurvey). Gesundheitswesen. 2003;65(6):393–401.

98. Takada M, Tsunekawa S, Hashimoto K, Inada T, Kim K-H, Oeda Y, et al. Analysis of the Effects and Causes of Driver Horn Use on the Acoustic Environment at Urban Intersections in Taiwan. Applied Sciences [Internet]. 2022; 12(12).

99. Banerjee D, Chakraborty SK, Bhattacharyya S, Gangopadhyay A. Evaluation and Analysis of Road Traffic Noise in Asansol: An Industrial Town of Eastern India. International Journal of Environmental Research and Public Health [Internet]. 2008; 5(3):[165-71 pp.].

100. Mahmood JA. What Do Car Horns Say? An Overview of the Non-Verbal Communication of Horn Honking. Open Journal of Social Sciences. 2021;9(8):375–88.

101. Raulin A-C, Doss SV, Trottier ZA, Ikezu TC, Bu G, Liu C-C. ApoE in Alzheimer’s disease: pathophysiology and therapeutic strategies. Molecular Neurodegeneration. 2022;17(1):72.

102. Slooter AJC, Cruts M, Kalmijn S, Hofman A, Breteler MMB, Van Broeckhoven C, et al. Risk Estimates of Dementia by Apolipoprotein E Genotypes From a Population-Based Incidence Study: The Rotterdam Study. Archives of Neurology. 1998;55(7):964–8.

