## Supplementary Materials for "Association of road traffic noise exposure with dementia or cognitive impairment – a systematic review of longitudinal cohort studies"

**Supplementary Material A: Search Strategy**

**Database: MEDLINE**

The literature search was conducted on MEDLINE using the Ovid platform on July 23, 2025. The search identified 985 records.

Ovid MEDLINE(R) ALL <1946 to July 23, 2025>

1	exp Noise, Transportation/ or exp Noise/	28785

2	((noise$ or sound$) adj pollution).ti,ab.	1273

3	((environmental or traffic or city or urban) adj2 noise$).ti,ab.	3748

4	1 or 2 or 3	31058

5	exp Dementia/ or exp Frontotemporal Dementia/ or exp Dementia, Vascular/	229541

6	exp Cognition Disorders/ or exp Cognitive Dysfunction/	130284

7	exp Cognition Disorders/ or exp Cognition/	327873

8	exp Alzheimer Disease/	134979

9	exp Neurocognitive Disorders/	346839

10	5 or 6 or 7 or 8 or 9	534786

11	(cognitive adj1 (impairment or deficit or decline or dysfunction$)).ti,ab.	146069

12	((frontotemporal or vascular) adj dementia).ti,ab.	17731

13	(alzheimer or (alzheimer adj disease$)).ti,ab.	33003

14	((cognitive or neurocognitive) adj disorder$).ti,ab.	11314

15	10 or 11 or 12 or 13 or 14	607269

16	4 and 15	1073

17	limit 16 to human	985

**Database: Embase**

The literature search was conducted on Embase using the Ovid platform on July 23, 2025. The search identified 2256 records.

Embase <1974 to 2025 July 23>

1	exp noise/ or exp traffic noise/ or exp noise pollution/	175055

2	((noise$ or sound$) adj pollution).ti,ab.	1500

3	((environmental or traffic or city or urban) adj2 noise$).ti,ab.	4567

4	1 or 2 or 3	176051

5	exp dementia/ or exp frontotemporal dementia/ or exp senile dementia/	515271

6	exp cognitive defect/	720414

7	exp Alzheimer disease/	287885

8	(cognitive adj1 (impairment or deficit or decline or dysfunction$)).ti,ab.	220741

9	((frontotemporal or vascular) adj dementia).ti,ab.	26364

10	(alzheimer or (alzheimer adj disease$)).ti,ab.	44643

11	((cognitive or neurogcognitive) adj disorder$).ti,ab.	10903

12	5 or 6 or 7 or 8 or 9 or 10 or 11	762256

13	4 and 12	2617

14	limit 13 to human	2256

**Database: EBSCOhost – GreenFile, CINAHL Plus**

The literature search was conducted on GreenFile and CINAHL Plus using the advanced search function on July 23, 2025. The search identified 55 records. The key words used in the search are as follows:

‘traffic noise pollution’ OR ‘road noise’ OR ‘traffic noise’ AND ‘dementia or alzheimers or cognitive impairment’ OR ‘cognitive decline’

**Supplementary Material B: Sample Data Extraction Form**

Date:

Extracted by:

Study:

1. Authors:
2. Year of publication:
3. Title:
4. Study design:
5. Age group:
6. Males/Females:
7. Duration of study:
8. Country:
9. Total participants at baseline:
10. Total participants in analysis:
11. Reasons for missing participants:
12. Exposure source:
13. Exposure measurement method:
14. Comparator:
15. Follow-up duration:
16. Outcome:
17. Outcome diagnosis method:
18. Outcome diagnostic criteria:
19. Statistical method used:
20. Unadjusted result (95% CI):
21. Adjusted result (95% CI):
22. Covariates adjusted for:
23. Key conclusions:

**Supplementary Materials Table C:**

Effect sizes and covariates adjusted for by each study included in the review.

| **Study** | **Exposure/Comparator** | **Result** | **Adjusted for** |
| --- | --- | --- | --- |
| Andersson et al. (2018) ^1^ | Leq 24h ≥ 55 dB reference < 55 dB | HR = 0.99 (95% CI: 0.62-1.59) | Baseline age. |
|  |  | HR = 0.97 (95% CI:0.58-1.60) | Baseline age, sex education, physical activity, smoking, BMI, waist-hip ratio, alcohol, ApoE4. |
|  |  | HR = 0.95 (95% CI:0.57-1.57) | Baseline age, sex education, physical activity, smoking, BMI, waist-hip ratio, alcohol, ApoE4, baseline medical history of diabetes, hypertension, stroke. |
| Cantuaria et al. (2021) ^2^ | Lnightmax  <40 dB  40-45 dB  45-50 dB  50-55 dB  ≥60 dB | HR = 1 (Reference)  HR = 1.11 (95% CI: 1.08-1.13) HR = 1.13 (95% CI: 1.11-1.15) HR = 1.13 (95% CI: 1.11-1.16) HR = 1.12 (95% CI: 1.10-1.15) | Age, sex, calendar year, civil status, income, region of origin, occupational status, proportion of high-quality green space, area-level socioeconomic variables: percent population with low income, only basic education, unemployed, with manual labour, single parents and with a criminal record, and mutual road traffic and railway noise adjustment. |
|  | Lnightmin  <40 dB  40-45 dB  45-50 dB  ≥50 dB | HR = 1 (Reference)  HR = 1.09 (95% CI: 1.08-1.11) HR = 1.07 (95% CI: 1.05-1.10) HR = 1.05 (95% CI: 1.01-1.09) | Age, sex, calendar year, civil status, income, region of origin, occupational status, proportion of high-quality green space, area-level socioeconomic variables: percent population with low income, only basic education, unemployed, with manual labour, single parents and with a criminal record, and mutual road traffic and railway noise adjustment. |
| Carey et al. (2018) ^3^ | Lnight  0-49.4  >49.4-49.6  >49.6-50.3  >50.3-53.8  >53.8 | HR = 1 (Reference)  HR = 1.05 (95% CI: 0.91-1.20)  HR = 1.04 (95% CI: 0.90-1.19)  HR = 1.00 (95% CI: 0.87-1.15)  HR = 1.09 (95% CI: 0.95-1.25) | Age, gender, ethnicity, smoking, alcohol consumption, BMI & IMD. |
| Ogurtsova et al. (2023) ^4^ | For every 10 dB increase in noise | Difference in standardised score = 0.125 (95% CI: -0.421, 0.67) | Nil |
|  |  | Difference in standardised score = 0.073 (95% CI: -0.474, 0.62) | Age, sex, iSES and nSES. |
|  |  | Difference in standardised score = 0.052 (95% CI: -0.496, 0.6) | Age, sex, iSES, nSES, alcohol consumption, BMI, diet, physical activity, smoking status, cumulative smoking, and environmental tobacco smoke exposure. |
| Yu et al. (2020) ^5^ | Lnight ≥ 55 dB reference < 55 dB | HR = 1.14 (95% CI: 0.79, 1.64) | Baseline age, gender, years of education. |
|  |  | HR = 1.13 (0.79, 1.63) | Baseline age, gender, years of education, occupation. |
|  |  | HR = 1.16 (0.80, 1.67) | Baseline age, gender, years of education, occupation, smoking status, alcohol consumption status, physical activity level. |
|  |  | HR = 1.18 (0.82, 1.71) | Baseline age, gender, years of education, occupation during most of life, smoking status, alcohol consumption status, physical activity level, neighbourhood socioeconomic status indicator, residential county. |
|  |  | HR = 1.20 (0.83, 1.73) | Baseline age, gender, years of education, occupation during most of life, smoking status, alcohol consumption status, physical activity level, neighbourhood socioeconomic status indicator, residential county, baseline Charlson index. |
|  |  | HR = 1.16 (0.80, 1.68) | Baseline age, gender, years of education, occupation during most of life, smoking status, alcohol consumption status, physical activity level, neighbourhood socioeconomic status indicator, residential county, baseline Charlson index, baseline cognition function and primary language. |
| Havyarimana et al. (2025) ^6^ | Lden 24h   <50 dB 50-55 dB  55-60 dB  ≥ 60 dB | Ref  HR = 0.98 (0.85, 1.13)  HR = 1.12 (0.93, 1.33)  HR = 1.03 (0.84, 1.26) | Age, sex, individual-level SES (education, household income, current employment status), area-level SES, cardiovascular risk score, PM_2.5_, green space. |
| Tuffier et al. (2024) ^7^ | Lden per-IQR increase | HR = 1.10 (1.02, 1.17) | Calendar year. |
|  |  | HR = 1.08 (1.01, 1.16) | Calendar year, BMI, smoking, alcohol consumption, employment and marital status, family income. |
|  |  | HR = 1.07 (0.99, 1.16) | Calendar year, BMI, smoking, alcohol consumption, employment and marital status, family income, area level covariates (municipality type, median wealth, frequencies of unemployment, inhabitants receiving financial aid, and high education), PM_2.5_. |
|  |  | HR = 1.02 (0.93, 1.11) | Calendar year, BMI, smoking, alcohol consumption, employment and marital status, family income, area level covariates (municipality type, median wealth, frequencies of unemployment, inhabitants receiving financial aid, and high education), PM_2.5_. |
| Wu et al. (2024) ^8^ | For every 10dB increase in Lden | HR = 1.02 (0.89, 1.17) | Age, sex, education, baseline year, birth year. |
|  |  | HR = 1.00 (0.87, 1.15) | Age, sex, education, baseline year, birth year, SES, smoking, physical activity, number of medications, neighbourhood household mean income, hearing loss. |
|  |  | HR = 0.98 (0.82, 1.16) | Age, sex, education, baseline year, birth year, SES, smoking, physical activity, number of medications, neighbourhood household mean income, hearing loss, other environmental pollution (PM_2.5_, green space and water space). |

*Abbreviations: Leq 24h = equivalent continuous sound level over 24 hours; dB = decibels; HR = hazard ratio; CI = confidence intervals; BMI = body mass index; ApoE4 = Apolipoprotein E4; Lnightmax = Lnight exposure at most exposed façade of residence; Lnightmin = Lnight exposure at least exposed façade of residence; Lnight = A-weighted, equivalent noise level at night; IMD = Indices of Multiple Deprivation; iSES = individual socioeconomic status; nSES = neighbourhood socioeconomic status.*

**Supplementary Materials Table D:**

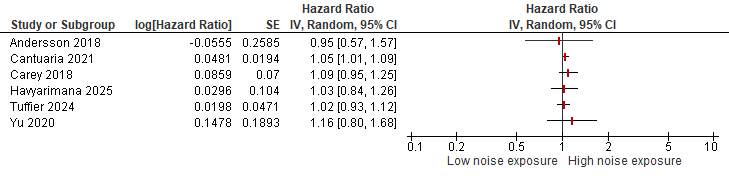

**Figure:** Forest plot of hazard ratios of dementia due to high noise exposure vs. low noise exposure using random effects model of eligible studies.
*SE = Standard error, IV = Inverse-variance, CI = confidence intervals.*

**References**

1.	Andersson J, Oudin A, Sundström A, Forsberg B, Adolfsson R, Nordin M. Road traffic noise, air pollution, and risk of dementia - results from the Betula project. Environ Res. 2018;166:334-9.

2.	Cantuaria ML, Waldorff FB, Wermuth L, Pedersen ER, Poulsen AH, Thacher JD, et al. Residential exposure to transportation noise in Denmark and incidence of dementia: national cohort study. BMJ. 2021;374:n1954.

3.	Carey IM, Anderson HR, Atkinson RW, Beevers SD, Cook DG, Strachan DP, et al. Are noise and air pollution related to the incidence of dementia? A cohort study in London, England. BMJ Open. 2018;8(9):e022404.

4.	Ogurtsova K, Soppa VJ, Weimar C, Jöckel K-H, Jokisch M, Hoffmann B. Association of long-term air pollution and ambient noise with cognitive decline in the Heinz Nixdorf Recall study. Environmental Pollution. 2023;331:121898.

5.	Yu Y, Mayeda ER, Paul KC, Lee E, Jerrett M, Su J, et al. Traffic-related Noise Exposure and Late-life Dementia and Cognitive Impairment in Mexican-Americans. Epidemiology. 2020;31(6):771-8.

6.	Havyarimana E, Gong X, Jephcote C, Johnson S, Suri S, Xie W, et al. Residential exposure to road and railway traffic noise and incidence of dementia: The UK Biobank cohort study. Environmental Research. 2025;279:121787.

7.	Tuffier S, Zhang J, Bergmann M, So R, Napolitano GM, Cole-Hunter T, et al. Long-term exposure to air pollution and road traffic noise and incidence of dementia in the Danish Nurse Cohort. Alzheimer's & Dementia. 2024;20(6):4080-91.

8.	Wu J, Grande G, Pyko A, Laukka EJ, Pershagen G, Ögren M, et al. Long-term exposure to transportation noise in relation to global cognitive decline and cognitive impairment: Results from a Swedish longitudinal cohort. Environment International. 2024;185:108572.
